# Mapping Individual Neuroanatomical Alterations to Schizophrenia Psychopathology with Normative Modeling

**DOI:** 10.64898/2026.03.31.26349848

**Authors:** Johanna Spaeth, Charlotte Fraza, Deniz Yilmaz, Lena Deller, BrainTrain Working Group, CDP Working Group, Genc Hasanaj, Marcel Kallweit, Maxim Korman, Mona Hussain, Emanuel Boudriot, Vladislav Yakimov, Joanna Moussiopoulou, Florian J. Raabe, Elias Wagner, Andrea Schmitt, Astrid Roeh, Peter Falkai, Daniel Keeser, Isabel Maurus, Lukas Roell

## Abstract

Schizophrenia spectrum disorders (SSDs) are clinically and neurobiologically heterogeneous. Normative modeling addresses heterogeneity of structural brain alterations by focusing on individual-level deviations, but their relation to clinical features requires further investigation. We mapped the relationship between individual gray matter volume (GMV) deviations and schizophrenia diagnosis and symptoms. Normative models of GMV were established using cross-sectional, T1-weighted magnetic resonance imaging data from a large, multi-site, healthy reference cohort (N = 7957). Deviations were derived for SSD patients (n = 379) and healthy controls (HCs, n = 149). We replicated that SSD is characterized by a more negative global deviation compared to HCs, yet extreme negative deviations exhibit substantial spatial heterogeneity. Regional deviations predicted diagnostic status with adequate performance (AUC = 0.79). A more negative deviation was associated with higher symptom severity and lower cognitive functioning in SSD. Negative deviations were scattered across the brain, with the largest alterations in the salience network. Our findings strengthen the potential of normative modeling to disentangle the heterogeneous underpinnings of SSD and provide further evidence for individualized structural deviations, particularly in the salience network, as promising markers of illness severity in SSDs.

---

Schizophrenia spectrum disorders (SSDs) are severe psychiatric conditions characterized by substantial interindividual differences in clinical phenotypes. Clinical heterogeneity is thought to be underpinned by neurobiological heterogeneity^1,2^. Although this notion is at the core of precision psychiatry frameworks such as the Research Domain Criteria^3^, case-control comparisons remain the dominant paradigm for identifying biomarkers. Obscuring variability within diagnostic categories, the reliance on group comparisons is a major barrier to understanding the pathophysiology of SSDs^4–8^.

Normative modeling is a statistical approach that accounts for heterogeneity by focusing on individual-level analyses. Analogous to pediatric growth curves for height and weight, normative models chart the relation between covariates and a (neuro)biological variable, such as gray matter volume (GMV), in a large reference population^6,7,9^. Unseen individuals can then be placed within this reference model to derive standardized measures of their deviation from the norm (the Z-score), resulting in an individualized deviation pattern across the brain.

By explicitly modeling interindividual variability rather than treating it as noise^7^, normative modeling provides a framework for disentangling biologically meaningful heterogeneity that underlies psychiatric diseases^10,11^. GMV is particularly well suited for this purpose, as it captures cumulative, trait-like neurobiological variation^12^, shows higher test-retest reliability compared with more state-dependent measures^13,14^, and may reflect relevant neuroanatomical alterations such as synapse loss^15^. To move beyond descriptive characterizations of structural heterogeneity towards a clinically informative framework, replication and extension of current approaches are essential.

Although parsing heterogeneity among clinical cohorts is a central objective of normative modeling, many studies have returned to a case-control perspective^10^. A common finding is lower overall centiles (greater negative deviations) in patients with SSDs compared to healthy controls (HCs) for GMV^16–19^ and other modalities^8,20–24^. However, this may not be representative of an individual given the highly heterogeneous loci of alterations among patients^19,22–26^. Further, group-level averages do not allow inferences as to whether Z-scores can predict diagnostic status at the individual-level^10^. Few studies have investigated machine learning (ML) classification of SSD patients vs. HCs using MRI-derived deviation features (e.g., cortical thickness, surface area or fractional anisotropy) with mixed results^20,27,28^. More evidence is needed to evaluate whether structural deviations can predict diagnostic status.

As classification still relies on predefined group labels, the usefulness of deviations depends on their association with clinical outcomes^6,7,10,29^. While Wolfers et al.^26^ found significant associations between global GMV deviation and psychotic symptom severity, this relationship was observed only across psychiatric groups and not within SSD specifically, and other studies have failed to detect significant links^22,30^. Associations between global deviation measures and cognitive functioning have been reported more consistently, although effect sizes vary^18,25–27,29^. Overall, characterizing associations of GMV deviations with clinical features requires further investigation.

Structural deviations are typically derived from specific brain regions, but psychiatric symptoms may arise from impaired network function rather than from dysfunction in isolated regions^19,31,32^. Aiming to resolve extensively documented heterogeneity of deviations at the regional level, it has been proposed that alterations converge onto common functional brain networks^19,33–35^. Accordingly, Janssen et al.^30^ found pronounced negative deviations in morphometric similarity within the default-mode network (DMN) in SSDs, whereas Segal et al.^19^ showed higher overlap of extreme negative deviations in GMV for almost all networks in schizophrenia compared to HCs. However, this effect was driven by a higher total deviation burden in patients, with only the salience/ventral attention network showing a specific impairment in schizophrenia^19^. The salience network plays an important role in SSDs, as disruptions closely map onto dopaminergic dysfunction^36,37^, are already evident in clinical high risk populations^38^, and as its function to assign importance to internal and external stimuli may explain positive and negative symptoms^39^. In summary, regionally heterogeneous structural deviations may map onto functional networks, but the specific networks mentioned differ between studies.

Building on previous research, the aim of this study is to map the relationship between individual-level GMV deviations and schizophrenia psychopathology. Aiming to replicate and extend current objectives in normative modeling, we ask the following research questions: (1) Is SSD associated with a negative deviation from the normative range of GMV, and do group-level abnormalities reflect individual patterns of deviation? (2) Can diagnostic status be predicted from deviation scores? (3) Are average GMV deviations associated with symptom severity, and (4) cognitive ability? (5) How do GMV alterations distribute across functional brain networks? We hypothesized that SSD patients would show a more negative average GMV deviation compared to HCs (H1), and that a more negative average deviation is correlated with higher symptom severity (H2) and worse cognitive functioning (H3) in patients.

Our work provides a comprehensive contribution to the field by addressing GMV deviations among patients and relative to controls, their links to symptomatology, and their spatial heterogeneity. From a methodological perspective, we introduce normative models for high-granularity, connectivity-based brain regions while addressing limitations of previous studies, such as insufficient sample sizes and restricted model fit evaluation criteria^10^. Leveraging a new clinical cohort, this study clarifies whether previous suggestions generalize across model implementations and samples.

## Methods

Analyses for this study were preregistered and can be accessed via the corresponding OSF repository (https://osf.io/46ev5/overview; https://osf.io/etdvu/overview?view_only=098deedc252e406697ddde6e9bfc2ee8). Figure 1 shows the study design.

**Figure 1.**
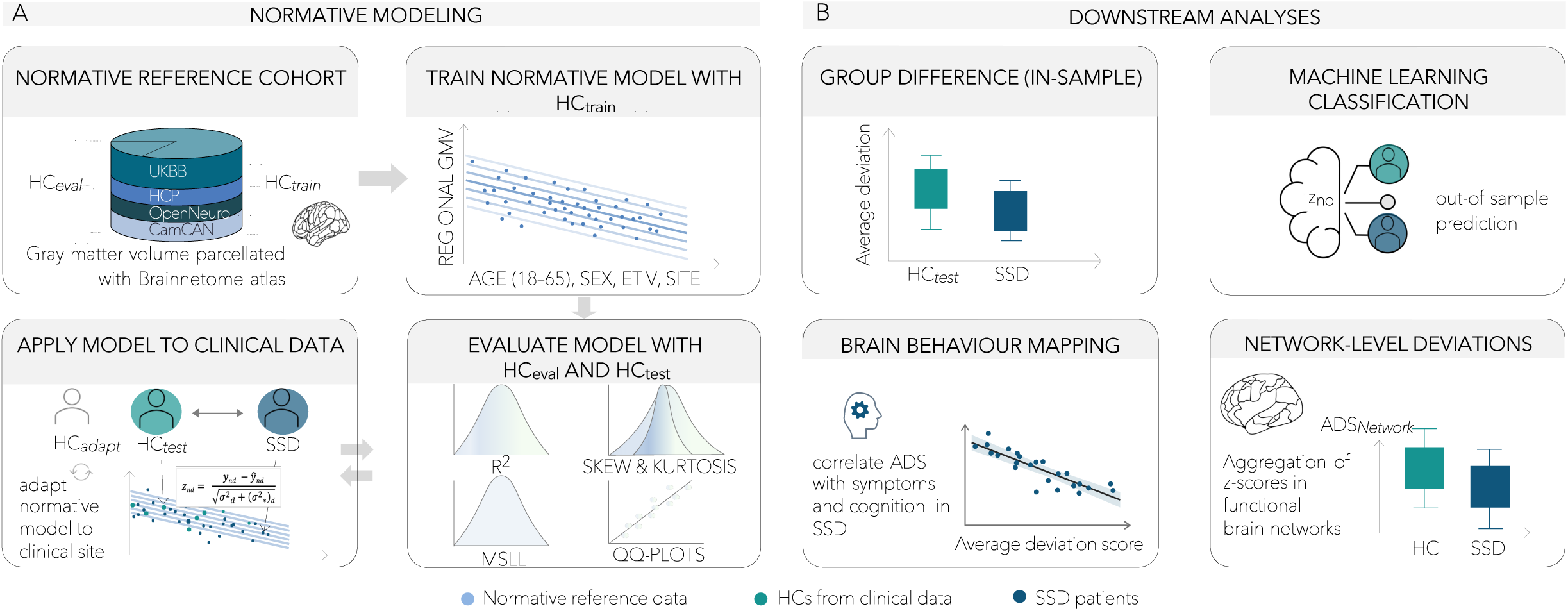
Study design and analysis pipeline. Panel A: The reference cohort was split into a training and evaluation (80/20) dataset. The training data were used to establish normative models, predicting GMV from age, sex, eTIV, and scanner site. Model fit was evaluated with R^2^, mean standardized log loss (MSLL), skew, and kurtosis in HC_eval_ and HC_test_. Models were adapted to the clinical site, upon which Z-scores for HC_test_ and SSD patients were derived. Panel B: Deviation scores from HC_test_ and SSD data were used in downstream analyses to compare average deviation scores (ADS), predict diagnostic status using machine learning, and correlate the ADS with cognitive ability and symptom severity in SSD. Z-scores were aggregated in functional brain networks and network ADS were compared within and between groups. eTIV: estimated total intracranial volume; GMV: gray matter volume; HC_eval_: healthy controls of the reference cohort used for model evaluation; HC_test_: healthy controls of the clinical cohort used for testing; SSD: schizophrenia spectrum disorder.

### Participants

The final analysis sample included HCs (n = 324) and SSD patients (n = 379) from six studies conducted at the Ludwig Maximilians University Hospital in Munich, here referred to as the *clinical datasets.* Research aims, study population, and ethics approval can be obtained from the original publications^40–42^ and Supplementary Material 1. For longitudinal studies, only baseline data were used, and we excluded subjects with missing data on any outcome variable (T1-weighted GMV) or covariate (age, sex, scanner site, eTIV) (see Supplementary Material 1 for sample sizes).

In brief, all participants were aged 18–65 years, had no history of neurological disease and no current abuse of illicit substances or cannabis. HCs had no current or past psychiatric disease. SSD samples comprised inpatients and outpatients with a diagnosis of schizophrenia, schizophreniform or schizoaffective disorder (according to ICD-10 or DSM-IV-TR/DSM-5) as confirmed by the Mini-International Neuropsychiatric Interview (M.I.N.I.^43^). Subjects with brief psychotic disorders, drug-induced psychotic disorder, bipolar disorders or any other severe psychiatric disease were excluded for the current project. Patients were treated with antipsychotic medication according to national treatment guidelines. All participants provided written informed consent, studies were approved by the local ethics committees and were conducted in accordance with the Declaration of Helsinki.

### MRI acquisition and processing

T1-weighted MRI scans were acquired with a magnetization-prepared rapid acquisition gradient-echo sequence at a 3-Tesla Siemens Magnetom Skyra scanner and 3-Tesla Siemens Magnetom Prisma scanner. Scanning protocols are summarized in Suppl. Table S2. T1-weighted images were first reoriented to standard anatomical space using *fslreorient2std*^44^. Intensity non-uniformity was corrected with N4BiasFieldCorrection^45^ and brain extraction was performed with the antsBrainExtraction workflow^46^. Tissue segmentation into gray matter (GM), white matter (WM), and cerebrospinal fluid (CSF) was carried out with FSL FAST. Global tissue volumes were computed from the partial-volume estimate (PVE) maps using *fslstats*. The brain-extracted T1w image was linearly registered to MNI152 space using FSL FLIRT. The resulting affine matrix was inverted with *convert_xfm*. Non-linear registration to the MNI template was then performed with FNIRT using the affine initialization and a membrane-energy regularization model. The deformation field was subsequently inverted using *invwarp*.

We used the Brainnetome atlas^47,48^ to parcellate the brain into 231 (thalamic regions combined, see Supplementary Material 1) cortical and subcortical GMV regions. It has shown to support robust estimations of functional connectomes^49^. Regions from the Brainnetome atlas were transformed from MNI space into native T1 space via *applywarp* with nearest-neighbor interpolation. Regional GM volumes were quantified for each atlas region using *fslstats* and the respective PVE map. Regional volumes were computed by weighting voxel counts by PVE intensities within each region of interest (ROI). All regional volume measures were saved as subject-level summary tables. This workflow is built on the Neuromodulation and Multimodal neuroimaging software (NAMNIs^50^; version 0.3). Estimated intracranial volume (eTIV) was extracted using the FreeSurfer (version 7.3.2) standard pipeline (*recon-all*).

### Scanner effect correction

Prior to modeling the clinical datasets as one fixed site in the normative models, we accounted for study-specific technical differences (e.g., head coil or scanner model) by employing a data harmonization approach. We provide details in Supplementary Material 1 and refer to Bayer et al.^51^ for a more general discussion on handling site effects. ComBat (http://enigma.ini.usc.edu/protocols/statisticalprotocols^52^) was used to correct for scanner-related variance in GMV, preserving (linear) effects of age, sex and eTIV. To avoid information leakage and generate unbiased estimates, ComBat parameters were first trained on a subset of held-out HCs (HC_adapt_) and subsequently applied to an HC sample unseen during training (HC_test_) and the SSD sample. Importantly, ComBat was never trained on patient data. As shown in Suppl. Figure S2 and Suppl. Figure S3, harmonization effectively removed study-related variance in both HC_adapt_ and the independent HC_test_ sample. Remaining between-study variation observed in the SSD sample is thus likely to reflect genuine differences in cohort characteristics (e.g., symptom severity; see Suppl. Figure S4 for study-specific PANSS scores).

### Quality control

Normative modeling requires rigorous data quality control to identify outliers due to technical errors (e.g., movement artifacts, segmentation errors) and extreme anatomical abnormalities while preserving biological variance^6^. We adopted a preregistered, stepwise outlier exclusion procedure combining data informed thresholds and expert visual inspection (as recommended^53^). A detailed description can be found in Supplementary Material 1. Overall, 70 subjects (0.62%) were excluded due to low data quality and the mean percentage of columns with missing values per participant was 0.12%. We subsequently imputed missing values with a random forest imputation technique (*missForestPredict*^54^). To avoid data leakage, we trained the imputation parameters on training sets (HC_adapt_; HC_train_) and applied them to unseen data (HC_test_/SSD; HC_eval_) (Supplementary Material 1).

### Establishing and evaluating normative models of GMV

Normative models were trained and evaluated on a large, healthy reference cohort derived from publicly available datasets. Only baseline scans were used, and subjects with a known diagnosis of any psychiatric disorder or missing data for any GMV value or covariate were excluded. We refer to the reference cohort as the nonclinical datasets, including de-identified data from the UK Biobank^55,56^, the Human Connectome Project (HCP) Young Adult^57^ and Aging^58^, the Cambridge Centre for Ageing and Neuroscience (CamCAN^59,60^) and datasets from OpenNeuro (https://openneuro.org/). A description of the datasets (including sample sizes) can be found in the original publications and in Supplementary Material 1. We split the nonclinical reference cohort into a training and test set (80/20) stratified by study site. The final sample size was N = 7957.

Models were trained using a Bayesian linear regression (BLR) from the PCNtoolkit (https://github.com/predictive-clinical-neuroscience/PCNtoolkit; version 0.28^6,8^), estimating regional GMV (i.e., independent models for each region) from age, sex (binary), scanner site (fixed effect) and eTIV. A cubic B-spline basis expansion with four evenly spaced knots was applied to model possible non-linear age effects while minimizing the risk of overfitting for centile curves in the restricted range (18–65 years). A fast numerical optimization algorithm (L-BFGS) was used as an optimization method. For details on the algorithm and implementation we refer to Fraza et al.^29^. A standard Gaussian noise distribution was modeled as the raw GMV data visually followed a normal distribution.

We evaluated the normative models in the independent test set of the normative reference cohort (HC_eval_) and in the clinical HC_test_ set. Following recommendations^10,29,61,62^, the model fit was measured with regard to the variance explained (R^2^), mean standardized log loss (MSLL, in HC_eval_) and the shape of the distribution (skew, kurtosis, QQ plots of the Z-scores).

### Applying clinical datasets to the normative models

HCs from the clinical datasets were split into a site adaptation set (HC_adapt_; n = 175) and a test set (HC_test_; n = 149), maintaining the distribution of age, sex and study (∼55/45 within-study-split). This step is necessary to calibrate the normative centile curves to the study-specific parameters while preventing data leakage. The remaining unbiased HC_test_ and SSD datasets were subsequently placed in the calibrated normative models to generate individual-level deviation scores (Z-scores). Z-scores quantify an individual’s deviation from the norm (per region) for a given set of covariates, with positive and negative values indicating higher or lower GMV values compared to the population median, respectively. As per previous work, Z-scores were calculated as

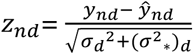

where the difference between the true (*y_nd_*) and the predicted (*y*#*_nd_*) GMV value for subject *n* in region *d* is normalized by the estimated predictive uncertainty. The Bayesian approach allows to incorporate two variance components, epistemic uncertainty ((*σ*^2^_∗_)*_d_*, induced by the data) and aleatoric uncertainty (*σ*^2^*_d_*, inherent to the underlying participant variability) (see e.g., Rutherford et al.^8^).

### Variables

#### Summary measures of deviation

As a primary outcome, the degree of deviation from the normative models was operationalized as the average deviation score (ADS), calculated by taking the mean of the regional Z-scores per individual (see ref. 28). For exploratory purposes, Z-scores were thresholded at Z > 1.96 and Z < −1.96 into supra- and infranormal Z-scores, marking extreme positive or negative percentiles (corresponding to uncorrected *p* < .05). The spatial overlap of extreme (negative) deviations was used to quantify heterogeneity. The individual-level percentage of infra- and supra-normal deviations relative to all Z-scores was calculated as a secondary outcome measure.

#### Symptom severity

Psychotic symptoms were measured with the total score of the Positive and Negative Syndrome Scale (PANSS^63^) ranging from 30–210 (see Supplementary Material 1 and Suppl. Figure S4 for study-specific PANSS distributions). Exploratorily, the negative and general subscales of the PANSS were used as outcome measures.

#### Cognitive performance

The Trail Making Test Version-B (TMT-B^64^) was used as a measure of cognitive performance, which predominantly reflects processing speed, a domain particularly impaired in SSD^65^, but also covers other cognitive domains (see Supplementary Material 1). Lower TMT-B scores reflect faster processing speed.

#### Network-level deviations

To calculate network-level deviations, we assigned GMV regions (excluding basal ganglia and thalamic regions) to a functional brain network of the Gordon atlas^66^ (Suppl. Table S4). We used the Network Correspondence Toolbox (NCT^67^) to generate Dice coefficients, which quantify the strength of connection between the region and the network. Regions were assigned to the network with the highest dice coefficient. To create a fair and more balanced comparison, we focused on those clinical datasets where studies included both HCs and SSD patients. A participant-level network deviation score (ADS_Network_) was calculated by taking the average Z-score across regions belonging to the network. Additionally, we quantified network-level proportions of extreme negative deviations.

### Statistical Analyses

Statistical Analyses were conducted with Python (version 3.11.5) and R^68^.

#### Confirmatory analyses

To test for a lower ADS in the SSD group compared to the HC group (H1), a one-sided t-test for independent samples was used (after Levene’s test suggested no significant difference in variances). Age, sex, scanning site, and eTIV were incorporated as predictors in the normative model used to derive the deviation scores and were therefore not included as additional covariates in the downstream analyses. For H2 and H3, assuming a significant negative correlation between the ADS and the total PANSS score (H2) and TMT-B score (H3) in the SSD group, a one-sided directional t-test for Spearman correlations (due to non-Gaussian data distributions) was implemented. All confirmatory hypotheses (H1–H3) were *p*-value corrected with the Bonferroni-Holm method controlling the family-wise error rate (FWER) at FWER = .05. For exploratory hypotheses, we focus on effect sizes.

#### Exploratory analyses

First, we investigated the confirmatory hypotheses with secondary outcome measures. The assumed higher percentage of infranormal (extreme negative) Z-scores in the SSD compared to the HC group was tested with a Mann-Whitney U directional test for independent samples. The correlation between the ADS and symptom severity (PANSS negative and general subscale) was analyzed with a directed t-test for the Spearman correlation coefficient. To investigate whether regional Z-scores can classify patients vs. controls at an individual-level, we conducted an ML benchmark experiment, using regional Z-scores as features. The performance of different algorithms, the Least Absolute Shrinkage and Selection Operator (LASSO^69^), Support Vector Machine (SVM^70^) and Random Forest (RF^71^) was compared in their ability to classify SSD vs. HC, using a 5 × repeated, nested 5-fold (2-fold inner loop) cross-validation (CV). Hyperparameter tuning was conducted within the inner loops; Z-scores were scaled within a preprocessing pipeline^72^. For a detailed description see Supplementary Material 1. The area under the curve (AUC) averaged across all outer CV folds was used as a primary performance measure.

To address the exploratory research question of network-level GMV alterations, we first identified networks with the most pronounced negative deviation in the patient group on a descriptive level. Individual-level network deviation scores were then compared between patients and controls using a regression approach (with network-level deviation scores as the dependent variables and diagnostic group (reference category: HC) as independent variable). Further exploring whether group differences result from global GMV reductions in SSD patients or additionally reflect network-specific alterations, we repeated the analysis, additionally controlling for a participant’s global network deviation (mean network-level ADS).

#### Sensitivity and supplemental analyses

To test the robustness of the results, we repeated the quality control procedure without the outlier cutoff decisions. We conducted several robustness analyses to investigate the effects of ComBat harmonization and repeated all analyses without harmonization using data from the largest included study. For the network-specific analysis, we additionally controlled for study in all analyses. The correlations were repeated using the raw data, controlling for the covariates in the sample. In supplemental analyses, we correlated the ADS with chlorpromazine equivalents (CPZ; according to the defined daily dose method^73^), illness duration and body mass index (BMI) where available. The ML benchmark experiment was repeated using raw GMV.

### Code and data availability

All programming scripts can be accessed on the OSF repository (https://osf.io/etdvu/overview?view_only=3a61e761f0bf4fb9b38baa15d7e15c61). Associated code from the PCNtoolkit can be found under https://github.com/predictive-clinical-neuroscience/braincharts. Links to publicly available data and access ways to controlled public data are described in Supplementary Material 1. Individual participant data from the clinical datasets cannot be shared publicly but are available upon reasonable request.

## Results

### Sample characteristics

Normative models were trained on HC_train_ = 6365 and the model fit was evaluated on HC_eval_ = 1592 healthy reference cohort individuals. The clinical sample included n = 324 HCs (54% site adaptation) and n = 379 SSD patients after exclusion of participants with missing data and scanning artifacts (see Table 1 for demographic and clinical characteristics in the collapsed SSD and HC cohort, Suppl. Table S1 for per-study results, Suppl. Table S3 for missing data exclusions and Suppl. Figure S1 for per-study age distributions). The SSD and HC samples differed significantly in age and sex composition, underscoring the importance of accounting for these variables in the normative deviation scores. The sample size is larger than suggested by the a priori power calculation for H1 (Supplementary Material 1).

**Table 1.** Descriptive statistics for clinical samples.

|  | HC<sub>adapt</sub> | HC<sub>test</sub> | SSD |  | *p* |
|---|---|---|---|---|---|
| N (% females) | 175 (45%) | 149 (44%) | 379 (31%) | *χ²*(1) = 7.26 | .007 |
| Age | 33 (12) | 34 (11) | 37 (11) | *t*(526) = 3.48 | < .001 |
| Clinical characteristics (only for patients) |  | M(SD) | range |  |  |
|  | PANSS total | 55 (16) | 30–104 |  |  |
|  | PANSS positive | 13 (5) | 7–29 |  |  |
|  | PANSS negative | 14 (6) | 7–32 |  |  |
|  | PANSS general | 28 (8) | 16–56 |  |  |
|  | TMT-B [sec] | 87 (52) | 25–366 |  |  |
|  | Illness duration [months] | 126 (112) | 0–516 |  |  |
|  | Age of onset | 27 (9) | 11–56 |  |  |
|  | CPZ | 406 (298) | 0–1963 |  |  |

*Note*. PANSS: Positive and Negative Syndrome Scale. TMT-B: Trail Making Test-B. CPZ = Chlorpromazine equivalents. N = 370 for PANSS total, n = 371 for PANSS negative and general. N = 266 for TMT-B. The statistical comparison is between HC<sub>test</sub> and SSD.

### Normative models of GMV

Normative models showed adequate performance in all measures. Using the HC_adapt_ data, the models could successfully be transferred to the clinical datasets that included sites unseen during training (HC_test_: EV > 0 for 87% of the models, |skew| < 1 and |kurtosis| < 3 for 98% of models; see Suppl. Figure S6). For any region, at least 86.6% (M = 97.9%, SD = 1.89) of HC_test_ individuals were placed within the normative range (−1.96 < Z < 1.96). For SSD patients, at least 85.2% (M = 96.01%, SD = 2.97) of deviation scores were within the normative range for any given region.

### Group differences in global GMV deviations

As hypothesized (H1, Figure 2A), we found a significantly lower average deviation score (ADS) in the SSD compared to the HC group (*t*(526) = −3.84, *p*_bonf_holm_ < .001; 95% CI ] −∞; −0.09]) with a small to medium effect size of *d* = −0.37 (95% CI [−0.56, −0.18]). Per-study ADS are visualized in Suppl. Figure S5. Overall, 76% of SSD patients exhibited at least one extreme negative deviation, compared with 59% of HCs. The proportion of participants showing at least one extreme positive deviation was 54% in both groups. SSD patients showed a greater burden of extreme negative deviations, with a median of 3 affected regions (IQR = 1–7), compared with 1 affected region in HCs (IQR = 0–2). Expressed as the proportion of affected regions across the whole brain, the median percentage of extreme negative deviations was 1.30% (M = 2.86%, SD = 4.84%) in patients and 0.43% (M = 1.24%, SD = 3.87%) in HCs. Comparing the percentage of extreme negative Z-scores as a secondary outcome measure, a Mann-Whitney U test supported the assumed higher percentage in patients (U = 37813, *p* < .001, *r*_rank-biserial_ = 0.34).

**Figure 2.**
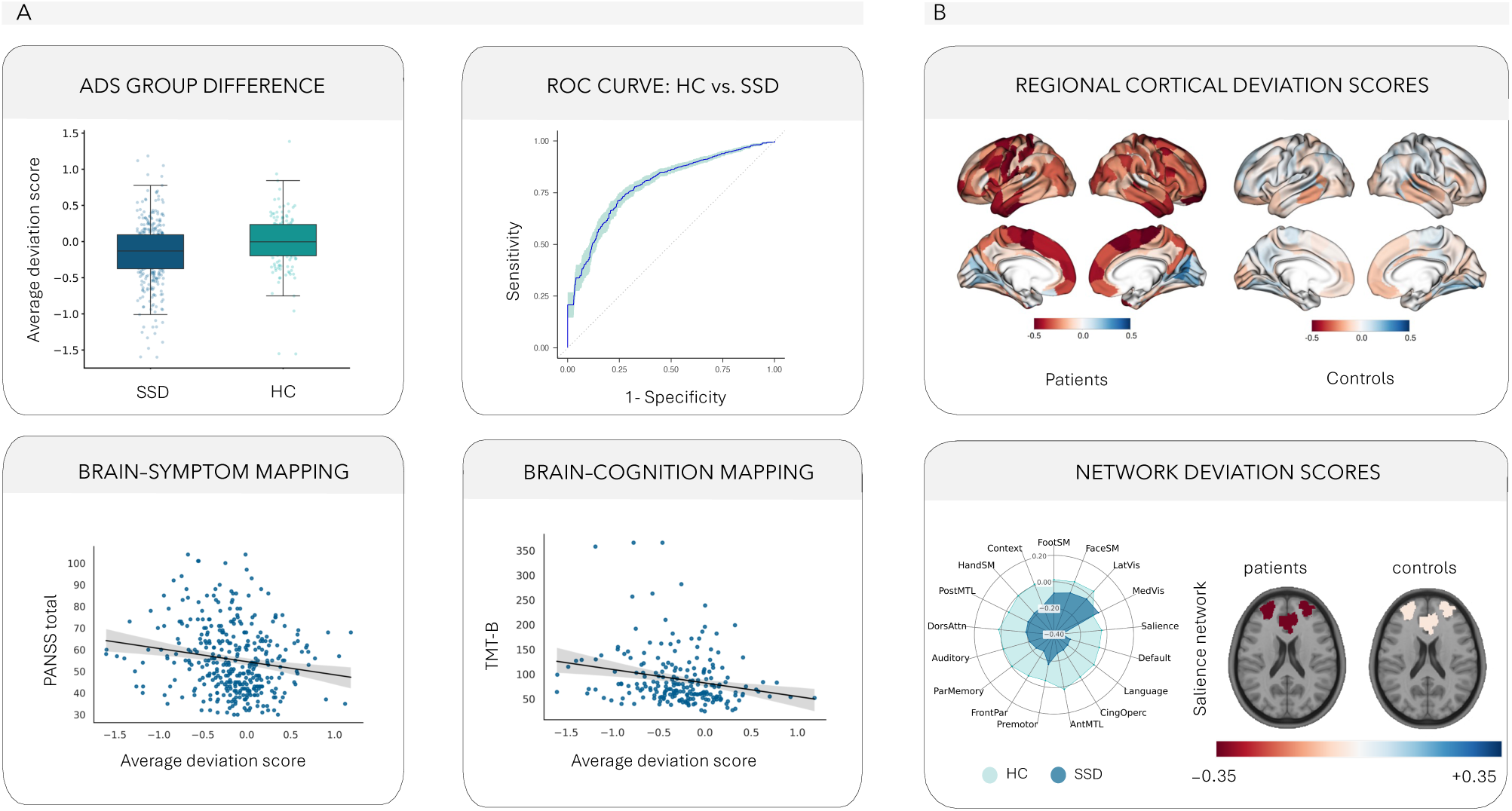
Global, regional and network-level gray matter deviations in SSD and their clinical associations. Panel A: Average deviation scores (ADS) between SSD patients and healthy controls (top left); receiver operating characteristic (ROC) curve for the classification of SSD vs. HC using a support vector machine (SVM) (top right). Association between the ADS and PANSS total (bottom left) and TMT-B completion time in seconds (bottom right) in SSD; statistics are Spearman correlations; lines show linear fits with 95% confidence intervals for visualization. Panel B: Mean regional Z-scores across the cortex in patients and controls (top); negative values (red) indicate lower gray matter volume than expected. Network-level ADS per group (bottom left; radial axis from −0.40 at the center to +0.20) and axial brain visualization of the mean salience network ADS per group (bottom right). Network abbreviations as in Suppl. Table S4. ADS: average deviation score; HC: healthy controls; PANSS: Positive and Negative Syndrome Scale; SSD: schizophrenia spectrum disorder; TMT-B: Trail Making Test Part B.

### Machine learning classification

An ML benchmark experiment was conducted to identify the best performing algorithm to classify SSD vs. HC from scaled regional Z-scores using a 5 × repeated, nested 5-fold CV resampling design. The SVM achieved the best performance with an AUC of 0.79 (SD = 0.05). Optimal hyperparameters were determined by retraining the SVM on the full dataset, yielding a radial basis function kernel with C = 0.681 and γ = 0.013. Figure 2A (top right) displays the receiver operating characteristic (ROC) curve for the SVM. Suppl. Table S5 shows secondary performance measures and results from supplemental analyses, where we included Z-scores from the HC_adapt_ dataset to create a larger and more balanced sample and achieved an AUC of 0.83 (optimal hyperparameters: radial basis function kernel, C = 11.121, γ = 0.009). Suppl. Table S6 shows results from the raw data ML benchmark.

### Associations of deviations with clinical features

A more negative ADS was correlated with higher symptom severity (PANSS total score; *ρ*(368) = −0.18, 95% CI [−0.28, −0.08], *p*_bonf_holm_ < .001) (H2; Figure 2A bottom left) and with worse cognitive functioning (TMT-B; *ρ*(264) = −0.23, 95% CI [−0.34, −0.11], *p*_bonf_holm_ < .001) (H3; Figure 2A bottom right). Exploratorily, a negative correlation was also present for the PANSS negative (*ρ*(369) = −0.20, 95% CI [−0.29, −0.10]) and PANSS general subscale (*ρ*(369) = −0.17, 95% CI [−0.27, −0.07]).

Results from all hypotheses (H1–H3) were robust to sensitivity analyses, where we did not apply outlier cutoff criteria (Supplementary Material 2). Likewise, the direction and magnitude of the effects did not change substantially when only using the non-harmonized data from participants in the largest included study (see Supplementary Material 2). In supplemental analyses, we repeated the correlation analyses with the raw features (mean GMV) regressing out the covariates in the sample, which yielded significant associations for cognitive function, but not for psychotic symptoms (Supplementary Material 2). We also report correlation analyses between the ADS and measures of medication, illness duration and BMI (Supplementary Material 2).

### Regional deviations and heterogeneity in SSD patients

Figure 2B (top) displays ADS for cortical regions. The most negative mean Z-score (−0.51) in patients was observed in the right paracentral lobule (a subregion of the superior frontal gyrus). In this region, the SSD sample also showed the greatest overlap of extreme negative (Z < −1.96) deviations (12%), whereas extreme positive (Z > 1.96) deviations were most pronounced (14%) in the left ventromedial putamen (a subregion of the basal ganglia).

### Network-level deviations

We assigned GMV regions to functional brain networks of the Gordon brain atlas^66^. For each individual, we calculated network-level ADS by taking the mean of the regional Z-scores belonging to the network. Patients exhibited the greatest negative deviation in the salience network (ADS_Salience_ = −0.34; Figure 2B bottom), followed by the DMN (ADS_DMN_ = −0.27). Comparing network-level ADS between SSD patients and HCs in a linear regression, the strongest group difference was observed for the salience network (β_group_ = −0.30, 95% CI [−0.41, −0.18], *p* < .001), followed by the anterior medial temporal lobe (aMTL) network (β_group_ = −0.28, 95% CI [−0.37, −0.19], *p* < .001) (see Suppl. Table S7 for network-wise group differences).

To investigate whether group differences exist beyond effects of total deviation burden, we controlled for an individual’s global network deviation burden in subsequent analyses. Again, the aMTL (β_group_ = −0.15, 95% CI [−0.22, −0.08], *p* < .001) and salience (β_group_ = −0.10, 95% CI [−0.15, −0.04], *p* < .001) networks showed the greatest negative (SSD < HC) group difference. Results did not change when additionally controlling for study in sensitivity analyses (Suppl. Table S8).

We additionally quantified proportions of extreme negative deviations at the network level. The highest overlap was observed in the aMTL network, where 35.6% of patients showed at least one extreme negative deviation (21.5% in HCs). For the average proportion of extreme negative deviations within each network, SSD patients showed the highest value in the DMN (3.7%), followed by the aMTL (3.5%) and salience (3.3%) networks.

Descriptively, the largest differences relative to HCs were observed in the DMN, aMTL and salience networks. In HCs, the average proportion of extreme negative deviations in these networks was 2.0%, 1.9% and 1.6%, respectively.

## Discussion

This study investigated whether individual neuroanatomical deviations from normative models map onto schizophrenia symptomatology. We report five main findings: (1) SSD diagnosis is associated with a global shift towards more negative GMV deviations from the norm compared to HCs, but extreme negative deviations showed only limited spatial overlap across patients. (2) Regional Z-scores classify patients vs. HCs with adequate predictive performance. (3) A more negative average deviation is associated with greater symptom severity and (4) lower cognitive functioning. (5) Spatially heterogeneous negative deviations are most pronounced in the salience network in SSD compared to HCs.

We extended previous evidence by establishing normative models for more fine-grained, connectivity-informed, whole-brain GMV regions and demonstrated links to clinical outcomes in a novel cohort. Our results confirm previous findings, which is particularly important as neuroimaging results can be highly unstable, with methodological choices such as preprocessing, analysis pipelines^49,74^ or atlas parcellation^75^ contributing to substantial variability. Hence, this study provides a key prerequisite for translating normative modeling into clinical practice.

As hypothesized, we found a more negative deviation in GMV for patients compared to HCs of small to medium effect size. Yet, this group-level effect did not translate into a common regional pattern, as extreme negative deviations were distributed heterogeneously across patients. We extended the group-level inference to predictive modeling in exploratory analyses, demonstrating that regional Z-scores classify patients vs. controls with adequate (AUC = 0.79) out-of-sample performance. Our results replicate previous studies that demonstrate in-sample^16–19^ and out-of-sample^20,27,28,76^ group differences in GMV deviations alongside marked interindividual spatial heterogeneity.

Different mechanisms may underlie structural alterations in SSD, including both atypical brain development and neurodegeneration^15^. Neurodevelopmental GMV loss may occur through the interaction of genetic (e.g., polygenic risk) and environmental (e.g., childhood maltreatment) risk factors, which contribute to synaptic dysfunction and glial-mediated synaptic pruning^15,77^. As the illness progresses, a feedback loop between psychotic symptoms and progressive synaptic and GMV loss may drive further neurodegeneration^77,78^. Importantly, recent studies have contradicted a simple neurodegenerative model in schizophrenia, demonstrating that deviations can normalize over time^16,79,80^, with group differences diminishing after 10 years^79^. On a side note, this aligns with our supplemental finding of a more positive average deviation being associated with a longer illness duration. Dimensionality of mental diseases thus not only exists across diagnostic categories but also along the course of illness, which limits the informativeness of the global group differences. Further, as deviations may not be pathological per se, often the case for positive or larger than expected GMV, it is essential to uncover links between deviations and psychopathology^10^.

We found that patients with a more negative deviation from the norm of GMV suffered from more severe total schizophrenia symptoms. The relation was stronger for negative symptoms, which were investigated exploratorily. Moreover, negative deviations were associated with worse cognitive functioning. Our results are in line with previous studies reporting associations between global deviations and psychopathology and cognition in SSD^26^, but contradict studies that have failed to do so^22,30^. More negative deviations have also been linked to higher depression severity^81^, suggesting at least partly transdiagnostic mechanisms. The finding that deviations showed stronger associations with symptoms than raw scores is also in accordance with previous studies^76^, emphasizing their added value in scenarios where population covariates mask underlying biological links^10^.

From a methodological perspective, variable results for brain-behavior associations may not only be attributable to sample characteristics or model choice, but also to the different operationalizations (e.g., mean deviation, percentage of extreme deviations) of deviations. Aligning with the idea of dimensionality, in this work we focused on the ADS, a location-independent summary measure that does not depend on thresholding. Although preferable in clinical practice, this global average might mask more differentiated associations between vulnerable regions and symptoms.

Indeed, some studies have reported associations of regional deviations with behavioral outcomes, both cross-sectionally^24,82,83^ and longitudinally^16,79,80^. On the one hand, symptoms may emerge from GMV loss. Howes and Onwordi^77^ propose that aberrant synaptic pruning leads to a cortical excitation-inhibition imbalance, resulting in cognitive and negative symptoms, as well as positive symptoms via dopamine dysregulation. On the other hand, stress that is associated with experiencing psychosis is speculated to contribute to further glial-mediated synaptic pruning, and hence GMV reductions^77^. When discussing underlying mechanisms, the modulatory role of antipsychotic medication must be considered. In supplemental analyses, a higher current medication dosage was associated with a more negative average deviation, consistent with evidence relating cumulative exposure to antipsychotics to progressive GMV loss in SSDs^84,85^. However, cross-sectionally, medication effects are difficult to disentangle from effects of illness severity. Notably, we observed associations with negative symptoms and cognitive functioning, that are typically only minimally influenced by antipsychotic treatment^86,87^. This argues against a strong medication-driven modulation of the deviation-symptom relationship in our sample, although this question requires further investigation in longitudinal studies. Taken together, our findings support the notion that individual structural alterations reflect illness severity in SSDs.

We found that GMV deviations were regionally scattered across the brain, with the maximum overlap of extreme negative deviations not exceeding 12% for any brain region. All brain networks exhibited negative deviations within the SSD sample. The salience network appeared to be most consistently ranked among the top deviating networks within and between groups, also when controlling for an individual’s global network deviation burden. Further, the DMN (within-group) and aMTL network (between-group) showed more pronounced negative deviations. Different parcellations and cutoffs used prevent a direct comparison, but the present pattern of regional alterations and heterogeneity aligns with previous findings from normative modeling^25,79^ and supports evidence for a widespread network impairment in SSD^19,88^, although other studies have suggested specific impairments, such as in the DMN^30^. Importantly, Segal et al.^19^ found only the salience network to be relatively affected beyond total deviation burden.

Despite a difference in the specific regional epicenters between individuals or subtypes^89,90^, their disruption may exert diaschisis-like effects on the entire network communication^89,91,92^. Network impairment could then explain common symptoms, such as impaired cognitive control or attention switching^19,23,30,33,34^. Along the course of illness, GMV alterations may propagate across the network^91^. Within the aberrant salience framework, early subcortical dysfunction in reward and salience processing is proposed to precede cortical abnormalities, with aberrant salience contributing to positive and negative symptoms^39^. Investigating mechanisms and behavioral consequences is beyond the scope of this study, but the exploratory analysis provides further support for network-level deviations in SSDs, particularly in the salience network.

### Limitations

Several limitations warrant consideration when interpreting the results. First, although including patients with different diagnoses from the psychosis spectrum, the study lacked a control group with other psychiatric conditions. Consequently, the group differences may not be specific to SSD, but may result from transdiagnostic factors, such as childhood trauma^81^ or health-related lifestyle^12^, although we did not find significant associations between average deviations and BMI in supplemental analyses. The limited availability of such variables prevented an assessment of their influence, including cumulative exposure to antipsychotic medication. Similarly, while we accounted for age, sex, and eTIV in the normative deviation scores, other factors, such as lower socioeconomic status and educational attainment, can contribute to reduced GMV in SSD^93–95^. Nevertheless, although (overall) structural deviations may not be disorder-specific or attributable to SSD *per se*, they appear to be especially pronounced in SSD^96^ and represent individual-level markers for a healthy brain^61^.

To achieve a large enough patient sample, we collapsed data from individual studies. Despite several robustness analyses, the employed harmonization approach is not without limitations, as scanner-related and biologically meaningful sources of between-study variance are difficult to fully disentangle. Further, whereas study site is modeled as a fixed effect within the normative modeling algorithm, different algorithms have been proposed that allow for a more nuanced handling of site effects^61,97^.

Further, although structural measures have shown to be informative when exploring deviations^11^, other modalities may provide additional insight, especially with regard to the network analyses. However, the connectivity-based parcellation of the Brainnetome atlas is conceptually well suited for a network-level perspective and offers an optimal basis for the integration of other modalities in the future.

Importantly, the current results are limited by the cross-sectional study design and do not allow for causal inferences. Future studies should include transdiagnostic samples, investigate longitudinal trajectories of deviations and their association with health-related factors, and combine multi-modal data sources.

Normative models are not exempt from the racial bias in neuroimaging^98,99^. Here, we emphasize that our goal was not to provide generalizable lifespan models, but to maximize precision for the deviations in the downstream analyses (i.e., by focusing on a restricted age range). Importantly, matching age distributions in the reference cohort has shown to substantially impact the robustness of deviations^100^. As such, despite leveraging a reference cohort with over 7000 individuals that is larger than common recommendations^27,99,101^, we refer to the extensively validated open-source normative models such as *CentileBrain*^27^ or the *braincharts* model^8^ for interested readers. Concluding with George E. P. Box’s aphorism “all models are wrong, but some are useful”^102^, the deviations should not be interpreted as absolute and *true* in a biological sense but can be useful due to their associations with disease characteristics.

## Conclusion

In conclusion, our findings demonstrate that despite a global shift towards more negative deviations in SSDs, negative deviations from the GMV norm are spatially heterogeneous among patients. We found significant deviation-symptom associations and provided evidence for network-level impairments. Our findings replicate and extend research avenues in normative modeling, emphasizing the value of individual-level analyses to disentangle the heterogeneous pathophysiology of SSDs.

## Acknowledgements

This research has been conducted using the UK Biobank Resource under Application Number 103216. Data were provided in part by the Human Connectome Project, WU-Minn Consortium (Principal Investigators: David Van Essen and Kamil Ugurbil; 1U54MH091657) funded by the 16 NIH Institutes and Centers that support the NIH Blueprint for Neuroscience Research; and by the McDonnell Center for Systems Neuroscience at Washington University. Data were further provided by the Human Connectome Project in Aging (HCP-A, U01AG052564), funded by the National Institute on Aging of the National Institutes of Health and by the McDonnell Center for Systems Neuroscience at Washington University in St. Louis. HCP-A data were obtained from the National Institute of Mental Health (NIMH) Data Archive (NDA). NDA is a collaborative informatics system created by the National Institutes of Health to provide a national resource to support and accelerate research in mental health. Dataset identifier: 2847 (collection ID). This manuscript reflects the views of the authors and may not reflect the opinions or views of the NIH or of the submitters submitting original data to NDA. Access to the Connectome Coordination Facility (CCF) permission group was last approved on July 24th, 2026 (DAR ID: 23525, OMB Control Number: 0925-0667). Data were further provided by the Cambridge Centre for Ageing and Neuroscience (CamCAN), for which funding was provided by the UK Biotechnology and Biological Sciences Research Council (grant number BB/H008217/1), together with support from the UK Medical Research Council and University of Cambridge, UK.

## Ethics declaration

Studies providing patient and healthy control data were approved by the ethics committee of the Faculty of Medicine, Ludwig-Maximilians-Universität München and were conducted in accordance with the Declaration of Helsinki. All participants provided written informed consent. Study-specific approval numbers are provided in Supplementary Material 1. Reference cohort data were obtained from publicly available or controlled-access resources (UK Biobank, Human Connectome Project, CamCAN, OpenNeuro), whose original studies were approved by the respective institutional review boards (see Supplementary Material 1), and were used in accordance with the respective data use agreements.

## Author contributions

JS and LR conceptualized the study. JS, LR and CF developed the methodology. JS, LR, VY, JM, FJR, EB, MKa and MKo curated the data. The BrainTrain Working Group (LD, PF, MH, IM, JM, LR, AS, JS, VY, DY) and the CDP Working Group (EB, PF, GH, MKa, DK, MKo, JM, FJR, LR, AS, EW, VY) collected the data (full lists of members are provided in the Supplementary Information). IM, JM, VY, EB, EW, DK and FJR provided resources. JS performed the formal analysis and created the visualizations. PF, AS and IM acquired funding. LR, IM, AR and PF supervised the project. JS wrote the original draft. All authors reviewed and edited the manuscript and approved the final version.

## Consortia

BrainTrain Working Group: Lena Deller^1^, Peter Falkai^1,7,11^, Mona Hussain^1^, Isabel Maurus^1,8^, Joanna Moussiopoulou^1,5^, Lukas Roell^1,5,7,12^, Andrea Schmitt^1,7,10,11^, Johanna Spaeth^1^, Vladislav Yakimov^1,8^ & Deniz Yilmaz^1,4,5^. A full list of members appears in the Supplementary Information.

CDP Working Group: Emanuel Boudriot^1,7^, Peter Falkai^1,7,11^, Genc Hasanaj^1,5,6^, Marcel Kallweit^1,5^, Daniel Keeser^1,5^, Maxim Korman^1,5^, Joanna Moussiopoulou^1,5^, Florian J. Raabe^1,7^, Lukas Roell^1,5,7,12^, Andrea Schmitt^1,7,10,11^, Elias Wagner^6,9^ & Vladislav Yakimov^1,8^. A full list of members appears in the Supplementary Information.

## References

1. Alnæs, D., Kaufmann, T., van der Meer, D., Córdova-Palomera, A., Rokicki, J., Moberget, T., et al. Brain Heterogeneity in Schizophrenia and Its Association With Polygenic Risk. JAMA Psychiatry 76, 739–748 (2019).

2. Voineskos, A. N., Jacobs, G. R. & Ameis, S. H. Neuroimaging Heterogeneity in Psychosis: Neurobiological Underpinnings and Opportunities for Prognostic and Therapeutic Innovation. Biol. Psychiatry 88, 95–102 (2020).

3. Insel, T., Cuthbert, B., Garvey, M., Heinssen, R., Pine, D. S., Quinn, K., et al. Research domain criteria (RDoC): toward a new classification framework for research on mental disorders. Am. J. Psychiatry 167, 748–751 (2010).

4. Brugger, S. P. & Howes, O. D. Heterogeneity and Homogeneity of Regional Brain Structure in Schizophrenia: A Meta-analysis. JAMA Psychiatry 74, 1104–1111 (2017).

5. Feczko, E., Miranda-Dominguez, O., Marr, M., Graham, A. M., Nigg, J. T. & Fair, D. A. The Heterogeneity Problem: Approaches to Identify Psychiatric Subtypes. Trends Cogn. Sci. 23, 584–601 (2019).

6. Marquand, A. F., Rezek, I., Buitelaar, J. & Beckmann, C. F. Understanding Heterogeneity in Clinical Cohorts Using Normative Models: Beyond Case-Control Studies. Biol. Psychiatry 80, 552–561 (2016).

7. Marquand, A. F., Kia, S. M., Zabihi, M., Wolfers, T., Buitelaar, J. K. & Beckmann, C. F. Conceptualizing mental disorders as deviations from normative functioning. Mol. Psychiatry 24, 1415–1424 (2019).

8. Rutherford, S., Fraza, C., Dinga, R., Kia, S. M., Wolfers, T., Zabihi, M., et al. Charting brain growth and aging at high spatial precision. eLife 11, e72904 (2022).

9. Marquand, A. F., Wolfers, T., Mennes, M., Buitelaar, J. & Beckmann, C. F. Beyond Lumping and Splitting: A Review of Computational Approaches for Stratifying Psychiatric Disorders. Biol. Psychiatry Cogn. Neurosci. Neuroimaging 1, 433–447 (2016).

10. Fraza, C., Rutherford, S., Bučková, B. R., Beckmann, C. F. & Marquand, A. F. The promise of quantifying individual risk for brain disorders through normative modeling, a narrative review. Neurosci. Biobehav. Rev. 176, 106284 (2025).

11. Fraza, C., Sønderby, I. E., Boen, R., Shi, Y., Beckmann, C. F. & Marquand, A. F. Unraveling the link between CNVs, cognition and individual neuroimaging deviation scores from a population-based reference cohort. Nat. Ment. Health 2, 1451–1463 (2024).

12. Pan, G., Zhang, Y., Kang, J.-J., Jiang, Y., Zhang, W., Ren, P., et al. Modifiable traits and genetic associations with grey matter volume in mid-to-late adulthood: a population-based study in the UK biobank. Npj Aging 11, 67 (2025).

13. Elliott, M. L., Knodt, A. R., Ireland, D., Morris, M. L., Poulton, R., Ramrakha, S., et al. What Is the Test-Retest Reliability of Common Task-Functional MRI Measures? New Empirical Evidence and a Meta-Analysis. Psychol. Sci. 31, 792–806 (2020).

14. Parsons, S., Brandmaier, A. M., Lindenberger, U. & Kievit, R. Longitudinal stability of cortical grey matter measures varies across brain regions, imaging metrics, and testing sites in the ABCD study. Imaging Neurosci. 2, imag–2–00086 (2024).

15. Howes, O. D., Cummings, C., Chapman, G. E. & Shatalina, E. Neuroimaging in schizophrenia: an overview of findings and their implications for synaptic changes. Neuropsychopharmacology 48, 151–167 (2023).

16. Alemán-Morillo, C., García-San-Martín, N., Bethlehem, R. A. I., Dorfschmidt, L., Alemany-Navarro, M., Segura, P., et al. Medication and atypical brain maturation in psychosis associated with long-term cognitive decline and symptom progression. Br. J. Psychiatry 1–11 (2025) doi:10.1192/bjp.2025.10482.

17. García-San-Martín, N., Bethlehem, R. A. I., Mihalik, A., Seidlitz, J., Sebenius, I., Alemán-Morillo, C., et al. Molecular and micro-architectural mapping of gray matter alterations in psychosis. Mol. Psychiatry 30, 1287–1296 (2025).

18. Kim, M., Leonardsen, E., Rutherford, S., Selbæk, G., Persson, K., Steen, N. E., et al. Mapping cerebellar anatomical heterogeneity in mental and neurological illnesses. Nat. Ment. Health 2, 1196–1207 (2024).

19. Segal, A., Parkes, L., Aquino, K., Kia, S. M., Wolfers, T., Franke, B., et al. Regional, circuit and network heterogeneity of brain abnormalities in psychiatric disorders. Nat. Neurosci. 26, 1613–1629 (2023).

20. Elad, D., Cetin-Karayumak, S., Zhang, F., Cho, K. I. K., Lyall, A. E., Seitz-Holland, J., et al. Improving the predictive potential of diffusion MRI in schizophrenia using normative models-Towards subject-level classification. Hum. Brain Mapp. 42, 4658–4670 (2021).

21. Haukvik, U. K., Wolfers, T., Tesli, N., Bell, C., Hjell, G., Fischer-Vieler, T., et al. Individual-level deviations from normative brain morphology in violence, psychosis, and psychopathy. Transl. Psychiatry 15, 118 (2025).

22. Lv, J., Di Biase, M., Cash, R. F. H., Cocchi, L., Cropley, V. L., Klauser, P., et al. Individual deviations from normative models of brain structure in a large cross-sectional schizophrenia cohort. Mol. Psychiatry 26, 3512–3523 (2021).

23. Segal, A., Smith, R. E., Chopra, S., Oldham, S., Parkes, L., Aquino, K., et al. Multiscale Heterogeneity of White Matter Morphometry in Psychiatric Disorders. Biol. Psychiatry Cogn. Neurosci. Neuroimaging 11, 1110–1122 (2026).

24. Worker, A., Berthet, P., Lawrence, A. J., Kia, S. M., Arango, C., Dinga, R., et al. Extreme deviations from the normative model reveal cortical heterogeneity and associations with negative symptom severity in first-episode psychosis from the OPTiMiSE and GAP studies. Transl. Psychiatry 13, 373 (2023).

25. Wolfers, T., Doan, N. T., Kaufmann, T., Alnæs, D., Moberget, T., Agartz, I., et al. Mapping the Heterogeneous Phenotype of Schizophrenia and Bipolar Disorder Using Normative Models. JAMA Psychiatry 75, 1146–1155 (2018).

26. Wolfers, T., Rokicki, J., Alnaes, D., Berthet, P., Agartz, I., Kia, S. M., et al. Replicating extensive brain structural heterogeneity in individuals with schizophrenia and bipolar disorder. Hum. Brain Mapp. 42, 2546–2555 (2021).

27. Ge, R., Yu, Y., Qi, Y. X., Fan, Y., Chen, S., Gao, C., et al. Normative modelling of brain morphometry across the lifespan with CentileBrain: algorithm benchmarking and model optimisation. *Lancet Digit*. Health 6, e211–e221 (2024).

28. ENIGMA Clinical High Risk for Psychosis Working Group. Normative Modeling of Brain Morphometry in Clinical High Risk for Psychosis. JAMA Psychiatry 81, 77–88 (2024).

29. Fraza, C. J., Dinga, R., Beckmann, C. F. & Marquand, A. F. Warped Bayesian linear regression for normative modelling of big data. NeuroImage 245, 118715 (2021).

30. Janssen, J., Guil Gallego, A., Díaz-Caneja, C. M., Gonzalez Lois, N., Janssen, N., González-Peñas, J., et al. Heterogeneity of morphometric similarity networks in health and schizophrenia. Schizophrenia 11, 70 (2025).

31. Goodkind, M., Eickhoff, S. B., Oathes, D. J., Jiang, Y., Chang, A., Jones-Hagata, L. B., et al. Identification of a Common Neurobiological Substrate for Mental Illness. JAMA Psychiatry 72, 305–315 (2015).

32. Sha, Z., Wager, T. D., Mechelli, A. & He, Y. Common Dysfunction of Large-Scale Neurocognitive Networks Across Psychiatric Disorders. Biol. Psychiatry 85, 379–388 (2019).

33. Ji, G.-J., Zalesky, A., Wang, Y., He, K., Wang, L., Du, R., et al. Linking Personalized Brain Atrophy to Schizophrenia Network and Treatment Response. Schizophr. Bull. 49, 43–52 (2023).

34. Liu, Z., Palaniyappan, L., Wu, X., Zhang, K., Du, J., Zhao, Q., et al. Resolving heterogeneity in schizophrenia through a novel systems approach to brain structure: individualized structural covariance network analysis. Mol. Psychiatry 26, 7719–7731 (2021).

35. Yang, G. J., Murray, J. D., Wang, X.-J., Glahn, D. C., Pearlson, G. D., Repovs, G., et al. Functional hierarchy underlies preferential connectivity disturbances in schizophrenia. Proc. Natl. Acad. Sci. U. S. A. 113, E219–228 (2016).

36. McCutcheon, R. A., Nour, M. M., Dahoun, T., Jauhar, S., Pepper, F., Expert, P., et al. Mesolimbic Dopamine Function Is Related to Salience Network Connectivity: An Integrative Positron Emission Tomography and Magnetic Resonance Study. Biol. Psychiatry 85, 368–378 (2019).

37. Winton-Brown, T. T., Fusar-Poli, P., Ungless, M. A. & Howes, O. D. Dopaminergic basis of salience dysregulation in psychosis. Trends Neurosci. 37, 85–94 (2014).

38. Del Fabro, L., Schmidt, A., Fortea, L., Delvecchio, G., D’Agostino, A., Radua, J., et al. Functional brain network dysfunctions in subjects at high-risk for psychosis: A meta-analysis of resting-state functional connectivity. Neurosci. Biobehav. Rev. 128, 90–101 (2021).

39. Kesby, J. P., Murray, G. K. & Knolle, F. Neural Circuitry of Salience and Reward Processing in Psychosis. Biol. Psychiatry Glob. Open Sci. 3, 33–46 (2023).

40. Krčmář, L., Jäger, I., Boudriot, E., Hanken, K., Gabriel, V., Melcher, J., et al. The multimodal Munich Clinical Deep Phenotyping study to bridge the translational gap in severe mental illness treatment research. Front. Psychiatry 14, 1179811 (2023).

41. Maurus, I., Hasan, A., Schmitt, A., Roeh, A., Keeser, D., Malchow, B., et al. Aerobic endurance training to improve cognition and enhance recovery in schizophrenia: design and methodology of a multicenter randomized controlled trial. Eur. Arch. Psychiatry Clin. Neurosci. 271, 315–324 (2021).

42. Moussiopoulou, J., Yakimov, V., Roell, L., Rauchmann, B.-S., Toth, H., Melcher, J., et al. Higher blood–brain barrier leakage in schizophrenia-spectrum disorders: A comparative dynamic contrast-enhanced magnetic resonance imaging study with healthy controls. Brain. Behav. Immun. 128, 256–265 (2025).

43. Sheehan, D. V., Lecrubier, Y., Sheehan, K. H., Amorim, P., Janavs, J., Weiller, E., et al. The Mini-International Neuropsychiatric Interview (M.I.N.I.): the development and validation of a structured diagnostic psychiatric interview for DSM-IV and ICD-10. J. Clin. Psychiatry 59 Suppl 20, 22–33;quiz 34-57 (1998).

44. Smith, S. M., Jenkinson, M., Woolrich, M. W., Beckmann, C. F., Behrens, T. E. J., Johansen-Berg, H., et al. Advances in functional and structural MR image analysis and implementation as FSL. NeuroImage 23, S208–S219 (2004).

45. Tustison, N. J., Avants, B. B., Cook, P. A., Zheng, Y., Egan, A., Yushkevich, P. A., et al. N4ITK: Improved N3 Bias Correction. IEEE Trans. Med. Imaging 29, 1310–1320 (2010).

46. Tustison, N. J., Cook, P. A., Holbrook, A. J., Johnson, H. J., Muschelli, J., Devenyi, G. A., et al. The ANTsX ecosystem for quantitative biological and medical imaging. Sci. Rep. 11, 9068 (2021).

47. Fan, L., Li, H., Zhuo, J., Zhang, Y., Wang, J., Chen, L., et al. The Human Brainnetome Atlas: A New Brain Atlas Based on Connectional Architecture. Cereb. Cortex 26, 3508–3526 (2016).

48. Jiang, T. Brainnetome: A new -ome to understand the brain and its disorders. NeuroImage 80, 263–272 (2013).

49. Luppi, A. I., Gellersen, H. M., Liu, Z.-Q., Peattie, A. R. D., Manktelow, A. E., Adapa, R., et al. Systematic evaluation of fMRI data-processing pipelines for consistent functional connectomics. Nat. Commun. 15, 4745 (2024).

50. Karali, T., Padberg, F., Kirsch, V., Stoecklein, S., Falkai, P. & Keeser, D. NAMNIs: Neuromodulation And Multimodal NeuroImaging software. Zenodo CERN Eur. Organ. Nucl. Res. https://doi.org/10.5281/zenodo.4547552 (2021) doi:10.5281/zenodo.4547552.

51. Bayer, J. M. M., Thompson, P. M., Ching, C. R. K., Liu, M., Chen, A., Panzenhagen, A. C., et al. Site effects how-to and when: An overview of retrospective techniques to accommodate site effects in multi-site neuroimaging analyses. Front. Neurol. 13, 923988 (2022).

52. Radua, J. combat.enigma: Fit and Apply ComBat, LMM, or Prescaling Harmonization for ENIGMA and Other Multisite MRI Data. (2024).

53. Rutherford, S. & Marquand, A. F. Normative Modeling with the Predictive Clinical Neuroscience Toolkit (PCNtoolkit). in Methods for Analyzing Large Neuroimaging Datasets (eds Whelan, R. & Lemaître, H.) 329–364 (Springer US, New York, NY, 2025). doi:10.1007/978-1-0716-4260-3_14.

54. Albu, E., Gao, S., Wynants, L. & Van Calster, B. missForestPredict—Missing data imputation for prediction settings. PLOS ONE 20, e0334125 (2025).

55. Miller, K. L., Alfaro-Almagro, F., Bangerter, N. K., Thomas, D. L., Yacoub, E., Xu, J., et al. Multimodal population brain imaging in the UK Biobank prospective epidemiological study. Nat. Neurosci. 19, 1523–1536 (2016).

56. Sudlow, C., Gallacher, J., Allen, N., Beral, V., Burton, P., Danesh, J., et al. UK Biobank: An Open Access Resource for Identifying the Causes of a Wide Range of Complex Diseases of Middle and Old Age. PLOS Med. 12, e1001779 (2015).

57. Van Essen, D. C., Smith, S. M., Barch, D. M., Behrens, T. E. J., Yacoub, E. & Ugurbil, K. The WU-Minn Human Connectome Project: An Overview. NeuroImage 80, 62–79 (2013).

58. Harms, M. P., Somerville, L. H., Ances, B. M., Andersson, J., Barch, D. M., Bastiani, M., et al. Extending the Human Connectome Project across ages: Imaging protocols for the Lifespan Development and Aging projects. NeuroImage 183, 972–984 (2018).

59. Shafto, M. A., Tyler, L. K., Dixon, M., Taylor, J. R., Rowe, J. B., Cusack, R., et al. The Cambridge Centre for Ageing and Neuroscience (Cam-CAN) study protocol: a cross-sectional, lifespan, multidisciplinary examination of healthy cognitive ageing. BMC Neurol. 14, 204 (2014).

60. Taylor, J. R., Williams, N., Cusack, R., Auer, T., Shafto, M. A., Dixon, M., et al. The Cambridge Centre for Ageing and Neuroscience (Cam-CAN) data repository: Structural and functional MRI, MEG, and cognitive data from a cross-sectional adult lifespan sample. NeuroImage 144, 262–269 (2017).

61. Dinga, R., Fraza, C. J., Bayer, J. M. M., Kia, S. M., Beckmann, C. F. & Marquand, A. F. Normative Modeling of Neuroimaging Data Using Generalized Additive Models of Location Scale and Shape. http://biorxiv.org/lookup/doi/10.1101/2021.06.14.448106 (2021) doi:10.1101/2021.06.14.448106.

62. Marquand, A., Rutherford, S. & Dinga, R. Fairly evaluating the performance of normative models. *Lancet Digit*. Health 6, e775 (2024).

63. Kay, S. R., Fiszbein, A. & Opler, L. A. The positive and negative syndrome scale (PANSS) for schizophrenia. Schizophr. Bull. 13, 261–276 (1987).

64. Reitan, R. M. & Wolfson, D. The Halstead-Reitan Neuropsychological Test Battery : Theory and Clinical Interpretation. (Neuropsychology Press, Tucson, Ariz, 1985).

65. Gebreegziabhere, Y., Habatmu, K., Mihretu, A., Cella, M. & Alem, A. Cognitive impairment in people with schizophrenia: an umbrella review. Eur. Arch. Psychiatry Clin. Neurosci. 272, 1139–1155 (2022).

66. Gordon, E. M., Laumann, T. O., Gilmore, A. W., Newbold, D. J., Greene, D. J., Berg, J. J., et al. Precision Functional Mapping of Individual Human Brains. Neuron 95, 791–807.e7 (2017).

67. Kong, R., Spreng, R. N., Xue, A., Betzel, R. F., Cohen, J. R., Damoiseaux, J. S., et al. A network correspondence toolbox for quantitative evaluation of novel neuroimaging results. Nat. Commun. 16, 2930 (2025).

68. R Core Team. R: A Language and Environment for Statistical Computing. (R Foundation for Statistical Computing, Vienna, Austria, 2024).

69. Tibshirani, R. Regression Shrinkage and Selection via the Lasso. J. R. Stat. Soc. Ser. B Methodol. 58, 267–288 (1996).

70. Cortes, C. & Vapnik, V. Support-vector networks. Mach. Learn. 20, 273–297 (1995).

71. Breiman, L. Random Forests. Mach. Learn. 45, 5–32 (2001).

72. Binder, M., Pfisterer, F., Lang, M., Schneider, L., Kotthoff, L. & Bischl, B. mlr3pipelines - Flexible Machine Learning Pipelines in R. J. Mach. Learn. Res. 22, 1–7 (2021).

73. Leucht, S., Samara, M., Heres, S. & Davis, J. M. Dose Equivalents for Antipsychotic Drugs: The DDD Method. Schizophr. Bull. 42, S90–S94 (2016).

74. Botvinik-Nezer, R., Holzmeister, F., Camerer, C. F., Dreber, A., Huber, J., Johannesson, M., et al. Variability in the analysis of a single neuroimaging dataset by many teams. Nature 582, 84–88 (2020).

75. Fürtjes, A. E., Cole, J. H., Couvy-Duchesne, B. & Ritchie, S. J. A quantified comparison of cortical atlases on the basis of trait morphometricity. Cortex 158, 110–126 (2023).

76. Rutherford, S., Barkema, P., Tso, I. F., Sripada, C., Beckmann, C. F., Ruhe, H. G., et al. Evidence for embracing normative modeling. eLife 12, e85082 (2023).

77. Howes, O. D. & Onwordi, E. C. The synaptic hypothesis of schizophrenia version III: a master mechanism. Mol. Psychiatry 28, 1843–1856 (2023).

78. Castro-de-Araujo, L. F., de Araujo, J. A. P., Morais Xavier, É. F. & Kanaan, R. A. A. Feedback-loop between psychotic symptoms and brain volume: A cross-lagged panel model study. J. Psychiatr. Res. 162, 150–155 (2023).

79. Berthet, P., Haatveit, B. C., Kjelkenes, R., Worker, A., Kia, S. M., Wolfers, T., et al. A 10-Year Longitudinal Study of Brain Cortical Thickness in People with First-Episode Psychosis Using Normative Models. Schizophr. Bull. 51, 95–107 (2025).

80. Bučková, B. R., Fraza, C., Rehák, R., Kolenič, M., Beckmann, C., Španiel, F., et al. Using normative models pre-trained on cross-sectional data to evaluate intra-individual longitudinal changes in neuroimaging data. eLife 13, (2025).

81. Bayer, J. M. M., Van Velzen, L. S., Pozzi, E., Davey, C., Han, L. K. M., Bauduin, S. E. E. C., et al. Dissecting heterogeneity in cortical thickness abnormalities in major depressive disorder: a large-scale ENIGMA MDD normative modelling study. Preprint at 10.1101/2025.03.17.643677 (2025).

82. Muñoz-Caracuel, M., Alemán-Morillo, C., García-San-Martín, N., Garrido-Torres, N., Alemany-Navarro, M., Bethlehem, R. A. I., et al. Predicting clinical and functional trajectories in individuals with first-episode psychosis by baseline deviations in grey matter volume. Br. J. Psychiatry J. Ment. Sci. 228, 419–427 (2026).

83. Remiszewski, N., Bryant, J. E., Rutherford, S. E., Marquand, A. F., Nelson, E., Askar, I., et al. Contrasting Case-Control and Normative Reference Approaches to Capture Clinically Relevant Structural Brain Abnormalities in Patients With First-Episode Psychosis Who Are Antipsychotic Naive. JAMA Psychiatry 79, 1133–1138 (2022).

84. Fusar-Poli, P., Smieskova, R., Kempton, M. J., Ho, B. C., Andreasen, N. C. & Borgwardt, S. Progressive brain changes in schizophrenia related to antipsychotic treatment? A meta-analysis of longitudinal MRI studies. Neurosci. Biobehav. Rev. 37, 1680–1691 (2013).

85. Vita, A., De Peri, L., Deste, G., Barlati, S. & Sacchetti, E. The Effect of Antipsychotic Treatment on Cortical Gray Matter Changes in Schizophrenia: Does the Class Matter? A Meta-analysis and Meta-regression of Longitudinal Magnetic Resonance Imaging Studies. Biol. Psychiatry 78, 403–412 (2015).

86. Feber, L., Peter, N. L., Chiocchia, V., Schneider-Thoma, J., Siafis, S., Bighelli, I., et al. Antipsychotic Drugs and Cognitive Function: A Systematic Review and Network Meta-Analysis. JAMA Psychiatry 82, 47–56 (2025).

87. Leucht, S., Leucht, C., Huhn, M., Chaimani, A., Mavridis, D., Helfer, B., et al. Sixty Years of Placebo-Controlled Antipsychotic Drug Trials in Acute Schizophrenia: Systematic Review, Bayesian Meta-Analysis, and Meta-Regression of Efficacy Predictors. Am. J. Psychiatry 174, 927–942 (2017).

88. Oliveira-Saraiva, D. & Ferreira, H. A. Normative model detects abnormal functional connectivity in psychiatric disorders. Front. Psychiatry 14, 1068397 (2023).

89. Fang, K., Wen, B., Niu, L., Wan, B. & Zhang, W. Higher brain structural heterogeneity in schizophrenia. Front. Psychiatry 13, (2022).

90. Shafiei, G., Markello, R. D., Makowski, C., Talpalaru, A., Kirschner, M., Devenyi, G. A., et al. Spatial Patterning of Tissue Volume Loss in Schizophrenia Reflects Brain Network Architecture. Biol. Psychiatry 87, 727–735 (2020).

91. Chopra, S., Segal, A., Oldham, S., Holmes, A., Sabaroedin, K., Orchard, E. R., et al. Network-Based Spreading of Gray Matter Changes Across Different Stages of Psychosis. JAMA Psychiatry 80, 1246–1257 (2023).

92. Pines, A. R., Frandsen, S. B., Drew, W., Meyer, G. M., Howard, C., Palm, S. T., et al. Mapping Lesions That Cause Psychosis to a Human Brain Circuit and Proposed Stimulation Target. JAMA Psychiatry 82, 368–378 (2025).

93. Crossley, N. A., Alliende, L. M., Czepielewski, L. S., Aceituno, D., Castañeda, C. P., Diaz, C., et al. The enduring gap in educational attainment in schizophrenia according to the past 50 years of published research: a systematic review and meta-analysis. Lancet Psychiatry 9, 565–573 (2022).

94. Zahra Rami, F., Kim, W.-S., Shen, J., Tsogt, U., Odkhuu, S., Cheraghi, S., et al. Differential effects of parental socioeconomic status on cortical thickness in patients with schizophrenia spectrum disorders and healthy controls. Neurosci. Lett. 804, 137239 (2023).

95. Hua, J. P. Y. & Dal Bon, K. Social Determinants of Health and Neurobiology Across the Schizophrenia Course: A Systematic Review. JAMA Psychiatry https://doi.org/10.1001/jamapsychiatry.2026.1312 (2026) doi:10.1001/jamapsychiatry.2026.1312.

96. Bethlehem, R. A. I., Seidlitz, J., White, S. R., Vogel, J. W., Anderson, K. M., Adamson, C., et al. Brain charts for the human lifespan. Nature 604, 525–533 (2022).

97. Bayer, J. M. M., Dinga, R., Kia, S. M., Kottaram, A. R., Wolfers, T., Lv, J., et al. Accommodating site variation in neuroimaging data using normative and hierarchical Bayesian models. NeuroImage 264, 119699 (2022).

98. Ge, R., Yu, Y., New, F., Haas, S. S., Sanford, N., Yu, K., et al. Empirical validation of race-neutral normative brain morphometry models across ethnoracially diverse populations. Proc. Natl. Acad. Sci. 123, e2521055123 (2026).

99. Rutherford, S., Wolfers, T., Fraza, C., Harnett, N. G., Beckmann, C., Ruhe, H. G., et al. To which reference class do you belong? Measuring racial fairness of reference classes with normative modeling. in Proceedings of the 9th Machine Learning for Healthcare Conference (PMLR, 2024).

100. Elleaume, C., Hebling Vieira, B., Floris, D. L., Langer, N. & the Australian Imaging Biomarkers and Lifestyle flagship study of ageing. Toward Robust Neuroanatomical Normative Models: Influence of Sample Size and Covariates Distributions. eLife 14, (2026).

101. Bozek, J., Griffanti, L., Lau, S. & Jenkinson, M. Normative models for neuroimaging markers: Impact of model selection, sample size and evaluation criteria. NeuroImage 268, 119864 (2023).

102. Box, G. E. P. & Draper, N. R. Empirical Model-Building and Response Surfaces. (Wiley, New York, 1987).

