## Supplementary Material for "Mapping Individual Neuroanatomical Alterations to Schizophrenia Psychopathology with Normative Modeling"

### Supplementary Material 1: Methods

#### Participants

The clinical datasets comprise the following individual studies: Clinical Deep Phenotyping (CDP[1]); Multimodal Imaging in Chronic Schizophrenia (MIMICSS[1]); Enhancing Schizophrenia Prevention and Recovery through Innovative Treatments (ESPRIT) C3[2]; IMPACT[3]; disorder-tailored transcranial direct current stimulation (tDCS) of the prefrontal cortex (MRSDC1); BrainTrain. Recruitment started in September 2014 (MIMICSS) and is still ongoing for one study (BrainTrain). We describe the studies in the following and refer to the original publications for further details.

***Clinical Deep Phenotyping (CDP) study***[1]

The CDP study was a naturalistic, single-center study conducted at the Department of Psychiatry and Psychotherapy, University Hospital of the Ludwig-Maximilians-University in Munich. The aim was to investigate the neurobiological underpinnings of clinically relevant schizophrenia subgroups. It included multilayer, transdiagnostic assessments and used a multimodal approach (including MRI assessments and clinical characterizations, neurocognitive assessments, multimodal brain imaging, electroencephalography, retinal anatomy and electrophysiology, genetic analysis, etc.). In- and outpatients with a diagnosis of SSD (schizophrenia, schizoaffective disorder, brief psychotic disorder, drug-induced psychosis and delusional disorder), bipolar disorder and major depressive disorder were recruited as well as HCs with no current or past psychiatric disorder. Patients were diagnosed (and HCs were screened) with the German Version of the Mini-International Neuropsychiatric Interview (M.I.N.I.[4] version 7.0.2) according to the DSM-5, text revision (DSM-5-TR), and ICD-10. Information about medication was obtained via medical records and self-reports. All participants were aged 18-65 years and had fluent German language skills. Exclusion criteria were any psychiatric disorder other than the above specified, pregnancy, existence of a concurrent clinically relevant neurological or neuropsychiatric disorder that affects the central nervous system or other severe somatic comorbidities, inability to provide written informed consent and relevant non-compliance that would interfere with the ability to participate in the study. Recruitment took place from July 14, 2021, until May 22, 2023. Patients were recruited from the University Hospital; HCs were recruited from the local area by online and in public advertisements. The study was approved by the local ethics committee at the LMU Munich, Germany (project no. 20-528 and 22-0035) and registered in the German Clinical Trials Register (ID: DRKS00024177). Data handling was embedded into the Munich Mental Health Biobank of the LMU Munich (ethics project no. 18-716) and used their approved data storage and data safety concept. The MRI assessment was performed with a Siemens Magnetom

Prisma 3T scanner and included T1-weighted magnetization-prepared rapid acquisition gradient echo (T1-MPRAGE) and other measurements. The Human Connectome Project (HCP) protocol was used for the MRI measurements.

***Multimodal Imaging in Chronic Schizophrenia Study (MIMICSS)***

The MIMICSS study was a pilot study based on the longitudinal PsyCourse study[5], for details see the supplementary material of the CDP study protocol publication[1]. It investigated imaging derived measures and their relation to cognitive deficits in chronic schizophrenia. The study was conducted at the University Hospital of the LMU Munich and took place from September 16, 2014, to April 3, 2018. Initially, 56 healthy controls (HC; 40 men and 16 women), 76 patients with schizophrenia (59 men and 17 women) and 22 unaffected relatives (UR; 6 men and 16 women) were included. Diagnosis was confirmed by two independent psychiatrists according to ICD-10 criteria, and the HC group was screened with the M.I.N.I. The mean age of patients with schizophrenia was 34.8 years (± 11.7 years); of HC 33.3 years (± 12.1 years); and of UR 42.7 years (± 17.0 years). All participants provided written informed consent, and the study was approved by the local ethics committee of the LMU Munich (project no. 17-13).

***Enhancing Schizophrenia Prevention and Recovery through Innovative Treatments (ESPRIT) C3 study[2].***

The ESPRIT C3 study was a multi-center, rater-blind, parallel-group, two-arm randomized controlled clinical trial investigating the effects of aerobic exercise vs. flexibility, balance and toning training in patients with post-acute schizophrenia. In- and outpatients aged 18–65 years with a primary diagnosis of schizophrenia according to DSM-IV-TR were included and assessed with the M.I.N.I. (version 6.0.0). Patients were in a post-acute disease stage and had to be treated with one or two antipsychotics in accordance with the current treatment guidelines (stable medication at least two weeks prior to study inclusion). Further inclusion criteria refer to the ability to provide consent, and females were required to have a negative pregnancy test (serum) at baseline and to use a reliable method of contraception. Exclusion criteria encompassed the inability to give informed consent, the presence of suicidality or being a risk to others, the presence of severe somatic or neurological comorbidities, history of assumption of relevant non-compliance that interferes with the ability to participate in a clinical trial, current drug abuse (positive urine drug screening, except benzodiazepines), lack of German language skills, and pregnancy or lactation. More details on the inclusion and exclusion criteria and baseline demographic information can be found in Table 2 of Maurus et al.[2] and in the corresponding study protocol publication[6]. The study was conducted at five sites in Germany (LMU Munich; Central Institute of Mental Health Mannheim; University Duesseldorf; University RWTH Aachen; Charité Berlin). For the current project, only data from the LMU Munich were used, to ensure comparability of the scanner site and protocol with the other studies. Further, only baseline data were used. The study was approved by the local ethics committees (project number 706–15, date 18.05.2016) and registered at ClinicalTrials.gov (NCT03466112) and in the German Clinical Trials Register (DRKS00009804).

***IMPACT study*[3].**

The study investigated blood-brain barrier leakage in SSDs, recruiting patients and HCs aged 18-60 years at the University Hospital of the LMU Munich. Patients with SSDs were in a post-acute disease stage and were treated as inpatients in the University Hospital. Inclusion criteria included age between 18 and 60 years and a diagnosis of SSD, according to DSM-5, assessed with the M.I.N.I., German version 7.0.2. All patients were treated with antipsychotic medication according to clinical guidelines. Exclusion criteria comprised current treatment with electroconvulsive therapy or non-invasive brain stimulation, coercive treatment, acute suicidality, any central nervous system (CNS) disorder, history of traumatic brain injury (TBI), severe somatic diseases, acute infections, current pregnancy or lactation, regular current drug abuse (in the past months), inability to provide informed consent, current participation in clinical trials and contraindication(s) to MRI or DCE-MRI. For the control group, age and sex-matched HCs with no past or current psychiatric disorder (as confirmed by the M.I.N.I.) were included. Exclusion criteria were equivalent to those described above. Recruitment for HCs was performed via announcements in digital channels (Hospital homepage, social media) and personal communication. Enrollment took place from June 2022 until October 2023. Assessments included MRI, contrast-enhanced MRI (DCE-MRI), clinical and cognitive assessments, CSF and blood acquisition. The study was approved by the local ethics committee of the Ludwig-Maximilians University Munich (project no. 21-1139) and conducted in accordance with the Declaration of Helsinki.

***Disorder-tailored Transcranial Direct Current Stimulation (tDCS) of the Prefrontal Cortex (MRSDC1).***

This double-blind placebo-controlled study was conducted at the University Hospital of the LMU Munich aiming to investigate the neurophysiological correlates of tDCS effects in patients with major depressive disorder and SSD compared to HCs. All subjects were randomized to receive either two tDCS or sham tDCS sessions (for details on the intervention see[7]). Inclusion criteria were age between 18–60 years and the ability to provide informed consent. Patients with schizophrenia were required to have a confirmed diagnosis according to ICD-10 criteria and be treated with stable antipsychotic medication at least one week prior to study inclusion. Exclusion criteria were contraindications for brain stimulation (e.g., history of brain surgery or severe brain injury), contraindications for MRI (e.g., metallic implants);

acute suicidality; treatment with electroconvulsive therapy in the present episode; treatment with deep brain stimulation or vagus nerve stimulation and/or any other intracranial implants; any other relevant psychiatric disorder; any relevant unstable medical condition and pregnancy. HCs were defined as having no current or past neurological or psychiatric disorder and were volunteers recruited from the local area. It was approved by the local ethics committee of the LMU Munich (project no. 493-14) and registered in the Clinical Trials Register (ClinicalTrials.gov; NCT02715128). For the current project, only baseline structural MRI data were used, which are independent of the intervention effects.

***BrainTrain study***

This ongoing study is conducted at the University Hospital of the LMU Munich, following a single-center, rater-blind, parallel-group, two-arm randomized controlled trial design. It investigates the effects of aerobic vs. flexibility, strength and balance training on hippocampal subfield volume changes in patients with SSDs. Inclusion criteria encompassed age between 18-65 years, a confirmed diagnosis of SSD according to ICD-10 as confirmed by the M.I.N.I, German Version 6.0.0. Patients were in and outpatients in a post-acute phase of the disorder, as indicated by a PANSS of ≤ 75 prior to study inclusion. A stable medication (at least two weeks) of no more than three antipsychotics was required prior to study inclusion. Female participants had to have a negative serum pregnancy test and agree to use a reliable method of contraception. Exclusion criteria encompassed the inability to provide written informed consent; acute suicidal ideation and risk to others; significant somatic or neurological comorbidities; other major psychiatric illnesses; a history of non-compliance likely to affect trial participation; a positive urine drug screen for illicit substances or cannabis (except benzodiazepines); insufficient German language proficiency; and pregnancy or lactation. Patients were randomized to either a three-month training of aerobic exercise (intervention) or flexibility, balance and strength training (control). MRI measurements and extensive clinical and cognitive testing were conducted at baseline, which were used in the current project. The study was approved by the local ethics committee (project no. 22-0921) and registered in the Clinical Trials Register (ClinicalTrials.gov; NCT05956327).

In the normative reference cohort, subjects from the following datasets were included:

***Cambridge Centre for Ageing and Neuroscience (CamCAN)*** *dataset*[8,9]

The CamCAN dataset covers multi-modal assessments in a population based cross-sectional sample across the adult lifespan, investigating age-related changes in cognition and brain structure and function and the neuronal underpinnings of cognitive aging. Funding was provided by the UK Biotechnology and Biological Sciences Research Council (grant number BB/H008217/1), together with support from the UK Medical Research Council and University of Cambridge, UK. Data collection and sharing for this project was provided by the CamCAN. For the current project, we used baseline structural MRI data and demographic covariates (age, sex) from non-psychiatric participants. Access approval was provided on September 17, 2024. All participants provided written informed consent, and experiments were performed in accordance with the relevant guidelines and regulations.

***UK Biobank***[10,11]

The UK (United Kingdom) Biobank is a very large, population-based prospective study, established to allow detailed investigations of the genetic and nongenetic determinants of the diseases of middle and old age. Recruitment of 500,000 participants and the collection of an unprecedented wealth of baseline data and samples were completed in 2010. The 500,000 participants were assessed between 2006 and 2010 in 22 assessment centers throughout the UK, covering a variety of different settings to provide socioeconomic and ethnic heterogeneity and urban–rural mix. A subset of participants was studied with multimodal imaging techniques (a subset of which used in this project, from the February 2020 release https://community.ukbiobank.ac.uk/hc/en-gb/articles/26655145866269-Past-data-releases). Ethics approval is granted by the North West Multi-centre Research Ethics Committee (latest renewal 2021), application number 103216.

***Human Connectome Project young adult (HCP-YA)***[12]

Data from the HCP-YA project is provided by the Human Connectome Project (HCP), WU-Minn Consortium (Principal Investigators: David Van Essen and Kamil Ugurbil; 1U54MH091657), funded by the 16 NIH Institutes and Centers that support the NIH Blueprint for Neuroscience Research; and by the McDonnell Center for Systems Neuroscience at Washington University. The HCP aimed to study and freely share data from 1200 young adult (ages 22-35) subjects from families with twins and non-twin siblings, using a protocol that includes structural and functional magnetic resonance imaging (MRI, fMRI), diffusion tensor imaging (dMRI) at 3 Tesla (3T) and behavioral and genetic testing (see https://www.humanconnectome.org/study/hcp-young-adult/document/1200-subjects-data-release). The 1200 Subjects Release (S1200) includes behavioral and 3T MR imaging data from 1206 healthy young adult participants (1113 with structural MR scans) collected in 2012-2015.

***Human Connectome Project-Aging (HCP-A)***[13,14]

The HCP-A dataset is a large, multi-modal, and freely available set of consistently acquired data for use by the scientific community to investigate and define normative developmental and aging related changes in the healthy human brain. Four acquisition sites used matched Siemens Prisma 3T MRI scanners with centralized quality control and data analysis. Data was acquired across multimodal imaging and behavioral domains with a focus on factors known to be altered in advanced aging. The HCP-A enrolled 1200+ healthy adults (ages 36-100+), with each study collecting longitudinal data in a subset of individuals at particular age ranges. It was funded under the auspices of the NIH Blueprint for Neuroscience Research, started in summer of 2016. Data and/or research tools used in the preparation of this manuscript were obtained from the National Institute of Mental Health (NIMH) Data Archive (NDA). NDA is a collaborative informatics system created by the National Institutes of Health to provide a national resource to support and accelerate research in mental health. Dataset identifier: 2847 (collection ID). Access to the Adolescent Brain Cognitive Development (ABCD)/Connectome Coordination Facility (CCF) permission group that contains the HCP-Aging dataset was last approved on July 24th, 2026 (DAR ID: 23525, OMB Control Number: 0925-0667). For the current project, baseline structural MRI data and relevant covariates (age, sex) were used.

***OpenNeuro datasets.***

For the current project, seven publicly available datasets from the OpenNeuro platform (https://openneuro.org/) were leveraged. For all datasets, we only used the baseline anatomical structural MRI measures and covariates (age, sex). Participants were recruited from the general population and were labelled as non-psychiatric. We describe the datasets in brief and refer to the OpenNeuro website for more detailed information about the study design and inclusion and exclusion criteria.

***Single Dose Intranasal Oxytocin Administration: Data from Healthy Younger and Older Adults (ds004725)***[15]. This dataset included data from generally healthy younger (n = 44, age range = 18-31 years, 48% female) and older adults (n = 43, age range = 63-81 years, 56% female) who self-administered a single dose (24 international units) of either intranasal oxytocin (OT) or a placebo (IND 100,860; NCT01823146). The study was conducted from August 2013 to October 2014. Healthy younger participants were recruited through the UF Psychology Department undergraduate participant pool, HealthStreet, handouts, and flyers. Healthy older participants were recruited through HealthStreet and UF participant registries. The study adopted a randomized, double-blind, between-subject design. The dataset consists of anatomical and functional resting-state neuroimaging scans acquired after nasal spray administration as well as study-specific phenotypic and demographic data. (only anatomical scans used here, not influenced by OT intake).

***Age differences in the neural basis of decision-making under uncertainty (ds004711)***[16]***.*** This dataset was conducted as part of the cross-sectional AgeRisk study, investigating the developmental trajectories of risk preference, impulsivity and self-control. The neuroimaging data contains structural scans from a sample of healthy human participants aged 16-81 years (n = 187). Data collection took place between 2016 and 2017 at the University Hospital in Basel, Switzerland.

***Listening task (ds004285)***[17]***.*** This dataset included 82 participants aged 19-81 with no history of neurological disease. They were randomly assigned to either a control (no motion feedback) or intervention (real-time motion feedback) group and performed a word repetition task. Baseline structural anatomical scans were used for the current project.

***National Institute of Mental Health (NIMH) Research Volunteer (RV) Data set (ds004215)***[18]***.*** This dataset is a comprehensive dataset characterizing healthy research volunteers. Assessments included clinical measurements, mood-related psychometrics, cognitive functioning neuropsychological tests, structural and functional MRI, diffusion tensor imaging and a comprehensive magnetoencephalography battery. To be eligible for the study, participants need to be medically healthy adults over 18 years of age with the ability to read, speak and understand English. All participants provided electronic informed consent for online pre-screening and written informed consent for all other procedures. Participants with a history of mental illness or suicidal or self-injury thoughts or behavior are excluded. Additional exclusion criteria include current illicit drug use, abnormal medical exam, and less than an 8th grade education or IQ below 70.

***Cognitive Control Theoretic Mechanisms of Real-time fMRI-Guided Neuromodulation (CTM) (ds003831)***[19]***.*** This dataset included healthy participants with no current psychiatric disorder as assessed by the SCID clinical interview for DSM-IV. Subjects were right-handed, native-born United States citizens, had no current use of psychotropic medication and had a negative illicit drug urine screen immediately prior to the MRI scan. The final participant sample included in Bush et al.[20] (n = 94, 65% female) had a mean age of 36.6 (SD = 13.8; range 18–64). Neuroimaging included T1-weighted baseline scans which are used for the current project.

***"Can we have a second helping? A replication study on the neurobiological mechanisms underlying self-control" (ds002643)***[21]***.*** Data from this study was collected as part of the research program Pilot Replication with project number 401.16.023, which is (partly) financed by the Dutch Research Council (NWO). It aimed to solidify the empirical evidence supporting self-control theory. To this end a preregistered replication study was conducted, where participants underwent functional magnetic resonance imaging while rating 50 food items on healthiness and tastiness and making choices about food consumption. A total of n = 80 participants (40 females, mean age 24.94 years; age range 18–43 years) who did not report having a history of psychiatric, neurological or metabolic illness completed the experiment conducted in the Netherlands. Baseline anatomical T1-weighted MRI scans were used for the current project.

***"Narratives: fMRI data for evaluating models of naturalistic language comprehension" (ds002345)***[22]***.*** The study collected data from n = 345 unique subjects participating in over 750 functional scans with accompanying anatomical data. All subjects reported having normal hearing and no history of neurological disorders. The task included auditory stimuli of 28 naturalistic spoken stories ranging from ~3 to ~56 minutes for a total of ~5 hours of unique audio stimuli. Data were collected over the course of ~seven years, from October 2011 to September 2018. The baseline anatomical T1-weighted MRI scans were used for the current project.

**Suppl. Figure S1.** Age distributions across datasets

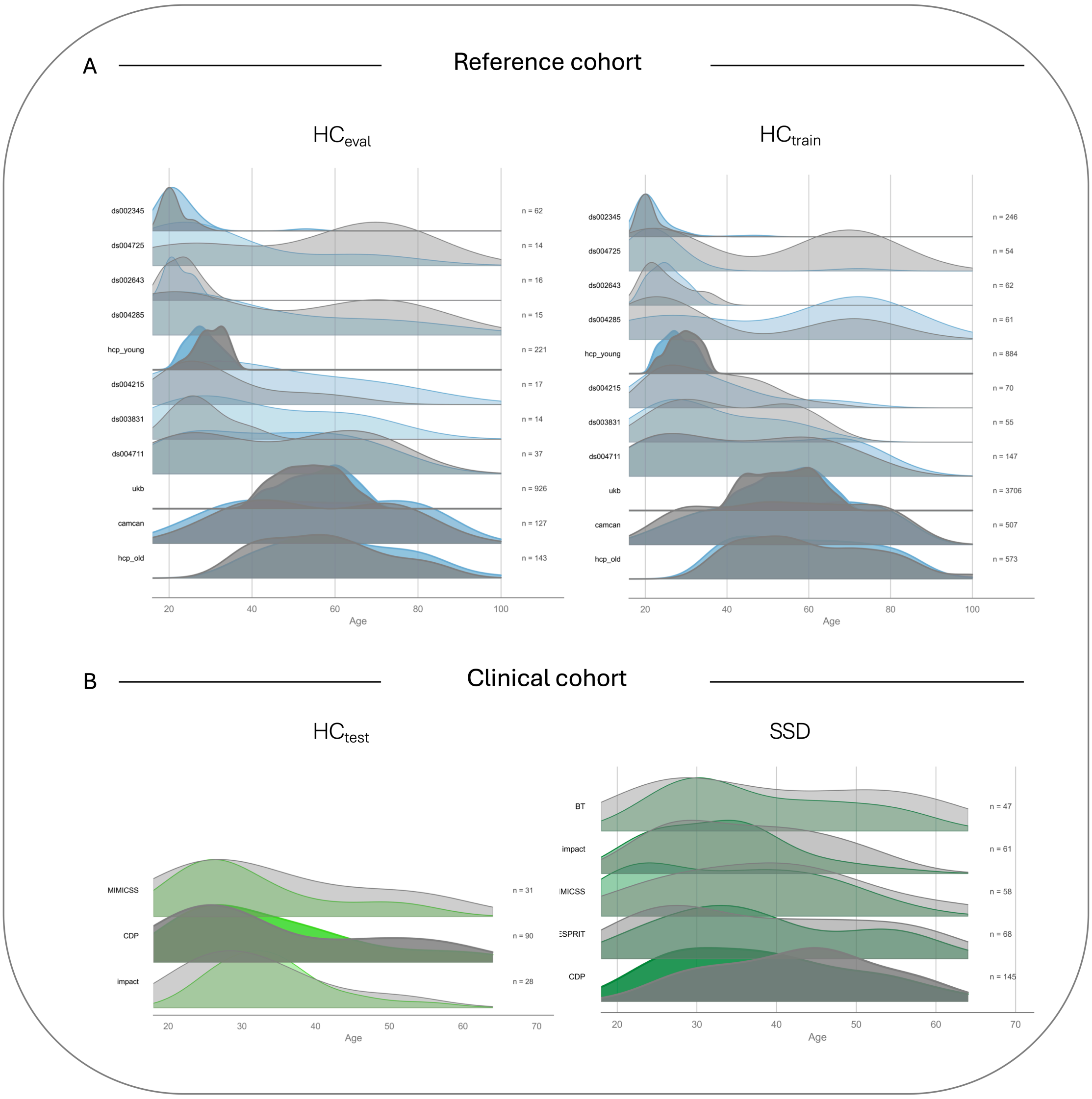


*Note*. Women are displayed in gray; men are displayed in color. Panel A: Per-site age distributions in the $HC_{eval}$ (left) and $HC_{train}$ (right) cohort. Panel B: Per-study age distributions in the $HC_{test}$ set (left) and SSD sample (right).

**Suppl. Table S1.** Descriptive Statistics for SSD patients separated by study

| | *M*(*SD*) [range] | | | | | *H*(4) | *p* | $\varepsilon^2$/V |
|---|---|---|---|---|---|---|---|---|
| | CDP | BT | IMPACT | ESPRIT | MIMICSS | | | |
| N | 145 | 47 | 61 | 68 | 58 | — | — | — |
| Age | 39 (11) [18-63] | 39 (12) [18-64] | 34 (9) [18-56] | 38 (12) [20-59] | 35 (11) [18-63] | 11.12 | .025 | .019 |
| Female, n (%) | 35 (24%) | 22 (47%) | 20 (33%) | 29 (43%) | 13 (22%) | $\chi^2(4)$ = 14.95 | .005 | .199 |
| Illness duration [months] | 149 (116) [0-516] | 129 (123) [2-480] | 99 (100) [0–432] | 125 (105) [2-440] | 96 (101) [0-408] | 14.81 | .005 | .029 |
| Age of onset | 27 (9) [15-56] | 27 (9) [15-55] | 26 (8) [11-50] | 28 (11) [15–56] | 27 (9) [15–52] | 0.31 | .989 | –.010 |
| CPZ | 339 (262) [0–1000] | 431 (369) [0–1560] | 420 (296) [0–1000] | 432 (225) [50–1071] | 511 (361) [0–1963] | 14.86 | .005 | .031 |
| PANSS | | | | | | | | |
| Total | 55 (17) [30-104] | 51 (10) [32–75] | 65 (15) [36–104] | 48 (11) [31-92] | 60 (15) [30–97] | 46.85 | <.001 | .117 |
| Positive | 13 (5) [7–29] | 11 (4) [7-24] | 16 (5) [7–29] | 11 (4) [7–18] | 13 (5) [7–23] | 35.70 | <.001 | .087 |
| Negative | 14 (6) [7–28] | 14 (5) [7–28] | 16 (6) [7–27] | 12 (4) [7–32] | 16 (5) [7–27] | 31.39 | <.001 | .075 |
| General | 29 (9) [16-56] | 25 (5) [17–35] | 33 (7) [19-50] | 25 (6) [16–45] | 30 (8) [16–49] | 44.72 | <.001 | .111 |
| TMT-B [sec] | 97 (64) [27–366] | 74 (31) [25–158] | 86 (55) [39-366] | 77 (34) [25-166] | 102 (65) [38–358] | 7.82 | .098 | .015 |

*Note*. Due to non-normal data distributions, between-study differences in continuous variables were assessed using Kruskal-Wallis tests, reported as *H*, *p* and $\varepsilon^2$. Differences in sex distributions were assessed using chi-square tests of independence, reported as $\chi^2$, *p*, and Cramér's V. PANSS: Positive and Negative Syndrome Scale. CPZ: Chlorpromazine equivalents. TMT-B: Trail Making Test-Version B. CDP: Clinical Deep Phenotyping. ESPRIT: Enhancing Schizophrenia Prevention and Recovery through Innovative Treatments. MIMICSS: Multimodal Imaging in Chronic Schizophrenia Study. BT: BrainTrain.

**Power calculation**

Leveraging several existing datasets, the sample size was determined by practical constraints. Nevertheless, a power analysis was conducted for the first hypothesis (H1) based on an estimated effect size reported in the literature using the *pwr* package[23]. Bethlehem et al.[24] established one of the largest normative models to date and compared mean centile scores for cortical and subcortical GMV between HCs and patients with schizophrenia. Effect sizes ranged from 0.11 (men, subcortical GMV) to 0.58 (women, cortical GMV), with an average between-group effect size of $d$ = 0.35. Conservatively assuming a between-group effect size of $d$ = 0.3 for the current analysis (directed t-test for independent samples), a significance level of $\alpha$ = .05 and a power of 1 - $\beta$ = .80, the required sample size would be N = 278 (n = 139 per group).

**Outlier exclusion/Quality control:**

We adopted a preregistered stepwise outlier exclusion procedure to identify subjects with technical artefacts (e.g., movement or segmentation errors). The aim was to identify implausible values, rather than outliers in the mathematical sense[25], preserving the biological variance of GMV. To this end, we first visualized data distributions of GMV for each region in all datasets. For the clinical datasets, we additionally visualized automatic image quality control metrics generated by MRIQC (version 24.0.0[26]). We marked subjects that showed very strong deviations from the distributions across regions and manually inspected their images (except for the UK Biobank data, for which manual checks were not feasible). Expert discussions guided the decision on inclusion/exclusion after manual data inspection. Subsequently, we mapped the relation between age (x-axis) and regional GMV (y-axis) for each dataset (separated for males and females). Visually guided outlier cutoffs indicating the maximum distance (in SD) from the sex- and study-specific regression line were established by consensus. For the clinical datasets, we chose a GMV value of >3SD away from the study- and sex-specific regression line as a criterion, whereas we chose a more lenient value of >3.5SD for the nonclinical datasets. We verified the cutoffs by manual quality checks for a subset of the data. To obtain a balance between accuracy and feasibility, we marked all values above the study-specific cutoff as missing and removed subjects with > 20% missing values (which was only the case for the UK Biobank data). Remaining missing values were imputed to maximize the available sample (as the PCNtoolkit requires complete data). As the thalamic regions appeared to be prone to segmentation errors (i.e., some areas had a value near zero, with the volume being assigned to another neighboring subregion), we decided on creating a summary variable by summing up the GMV values of all thalamic subregions (for an in-depth normative modeling analysis of thalamic nuclear volumes see Young et al.[27]).

**Suppl. Table S2.** Scanning protocols for clinical datasets

| Site/Study | Head coil | resolution | TR | TE | FA | slices |
|---|---|---|---|---|---|---|
| CDP/BrainTrain | 32-channel | 0.8 × 0.8 × 0.8 mm³ | 2500 | 2.2 | 8° | 208 |
| IMPACT | 64-channel | 0.8 × 0.8 × 0.8 mm³ | 2500 | 2.2 | 8° | 208 |
| ESPRIT/ MIMICSS/ MRSDC1 | 32-channel | 0.8 × 0.8 × 0.8 mm³ | 2060 | 2.2 | 12° | 256 |

*Note*. All T1-weighted images were acquired with a magnetization-prepared rapid gradient-echo (MPRAGE) sequence. TR: repetition time; TE: echo time; FA: flip angle; Slices: number of acquired slices.

**Missing data imputation**

To generate unbiased imputed values, we used a random forest imputation technique (*missForestPredict*[28]). For the clinical datasets, we trained the imputation parameters on the (site-harmonized) held-out $HC_{adapt}$ data and applied the parameters to the unseen (site-harmonized) $HC_{test}$ and SSD data. For the nonclinical datasets, we trained the imputation parameters on the training reference cohort $HC_{train}$ (separately for each dataset) and subsequently applied them to the respective datasets in the test set ($HC_{eval}$).

**Suppl. Table S3**. Sample sizes, missing data, and quality-control exclusions across datasets

| Dataset | Original N | N after QC | N after removal of missing data | % NA per participant (M, range) | % NA per GM column (M, range) |
|---|---|---|---|---|---|
| UKB | 7064 | 7032 | 4632 | 0.10% (0.0–19.48%) | 0.10% (0.0–0.45%) |
| HCP-YA | 1109 | 1109 | 1105 | 0.09% (0.0–4.33%) | 0.09% (0.0–0.81%) |
| HCP Aging | 725 | 722 | 716 | 0.12% (0.0–2.60%) | 0.12% (0.0–0.70%) |
| CamCAN | 640 | 637 | 634 | 0.12% (0.0–5.63%) | 0.12% (0.0–0.79%) |

| | | | | | |
|---|---|---|---|---|---|
| ds004725 | 87 | 84 | 68 | 0.03% (0.0–0.43%) | 0.03% (0.0–1.47%) |
| ds004711 | 186 | 185 | 184 | 0.08% (0.0–4.33%) | 0.08% (0.0–1.63%) |
| ds002345 | 340 | 340 | 308 | 0.09% (0.0–6.06%) | 0.09% (0.0–1.30%) |
| ds002643 | 80 | 78 | 78 | 0.02% (0.0–0.43%) | 0.02% (0.0–1.28%) |
| ds003831 | 73 | 69 | 69 | 0.02% (0.0–0.43%) | 0.02% (0.0–1.45%) |
| ds004215 | 149 | 146 | 87 | 0.05% (0.0–2.60%) | 0.05% (0.0–1.15%) |
| ds004285 | 78 | 76 | 76 | 0.03% (0.0–2.16%) | 0.03% (0.0–1.32%) |
| ESPRIT | 72 | 72 | 68 | 0.33% (0.0–6.49%) | 0.33% (0.0–2.94%) |
| CDP | 331 | 328 | 326 | 0.35% (0.0–8.66%) | 0.35% (0.0–1.84%) |
| MIMICSS | 124 | 116 | 109 | 0.23% (0.0–4.33%) | 0.23% (0.0–2.75%) |
| IMPACT | 132 | 126 | 117 | 0.17% (0.0–3.46%) | 0.17% (0.0–2.56%) |
| MRSDC1 | 36 | 36 | 36 | 0.01% (0.0–0.43%) | 0.01% (0.0–2.78%) |
| BrainTrain | 47 | 47 | 47 | 0.17% (0.0–1.73%) | 0.17% (0.0–4.26%) |

*Note.* From the initial datasets, subjects were excluded due to data quality and missing data. For the remaining subjects, GM values above outlier cutoffs were set to missing (NA). NA: not available. GM: gray matter. UKB: United Kingdom Biobank. HCP-YA: Human Connectome Project-Young Adult. CamCAN: Cambridge Centre for Ageing and Neuroscience. ESPRIT: Enhancing Schizophrenia Prevention and Recovery through Innovative Treatments. CDP: Clinical Deep Phenotyping. MIMICSS: Multimodal Imaging in Chronic Schizophrenia Study. MRSDC1: Disorder-tailored Transcranial Direct Current Stimulation (tDCS) of the Prefrontal Cortex.

**Data harmonization**

We follow recommendations[29] and model site as a fixed intercept in the normative models. Data harmonization strategies can become problematic when samples differ in variables unrelated to scanner effects, including various unknown demographic characteristics[30]. HCs included in the clinical datasets were recruited from the same overarching cohort (i.e., from the same university hospital, within a comparable time frame, and using identical recruitment procedures). Consequently, between-study differences in MRI measures are more plausibly attributable to differences in scanners, acquisition protocols, and head coils than to systematic biological differences between HC participants. We used the ComBat algorithm from the ENIGMA consortium[31] to harmonize scanner-related effects, preserving effects of age, sex and eTIV. Guided by visual inspection of age-related trends of GMV in these samples, we decided to model linear age effects to reduce the complexity of the models and the risk of overfitting. A further advantage of this approach is the implementation of a train-test split procedure (currently not available for non-linear ComBat GAM versions). This allowed us to train the harmonization parameters in a training set ($HC_{adapt}$) and apply them to independent test sets (SSD and $HC_{test}$). As such, data leakage from the harmonization step is prevented and fair comparisons can be made between the SSD and the $HC_{test}$ sample in downstream analyses.

To evaluate the effectiveness of ComBat harmonization in removing scanner-related variance, we quantified site effects before and after harmonization separately in the ComBat training dataset ($HC_{adapt}$), the independent ComBat test dataset ($HC_{test}$), and the SSD patient dataset.

For each of the 231 gray matter regions of interest (ROIs), site effects were evaluated using nested-model analyses of variance (ANOVAs). Specifically, a baseline linear model including age, sex, and eTIV was compared with a second model additionally including site as a fixed effect. The incremental variance explained by site was assessed using an extra sum-of-squares F-test obtained from the ANOVA comparison of the two nested models. This procedure was performed separately for the raw and ComBat harmonized data. P-values were adjusted for multiple comparisons across ROIs using the Bonferroni-Holm procedure.

As shown in Suppl. Figure S2, site effects were widespread before harmonization. In the ComBat training dataset, 210 of 231 ROIs (90.9%) showed significant site effects after Bonferroni-Holm correction. Similarly, significant site effects were observed in 200 of 231 ROIs (86.6%) in the independent test HC dataset and in 213 of 231 ROIs (92.2%) in the patient dataset. Following ComBat harmonization, no ROI exhibited a (corrected) significant site effect in either the training or test control datasets. Consistent with this finding, the median ROI-specific F statistic decreased from 46.46 to 0.01 in the training dataset and from 58.22

to 1.25 in the test dataset. In the patient dataset, the number of ROIs exhibiting significant site effects was reduced from 213 (92.2%) to 137 (59.3%), with the median F statistic decreasing from 118.13 to 10.70.

These findings indicate that ComBat effectively removed scanner-related variance in both the training and independent HC test datasets. Importantly, the harmonization parameters generalized to previously unseen HC data, suggesting that scanner effects were successfully estimated and removed without overfitting to the training sample. In contrast, between-study variability remained in the patient dataset following harmonization, which is expected due to genuine cohort differences, such as illness severity.

**Suppl. Figure S2.** Regions with significant site effects before and after ComBat harmonization separated by dataset

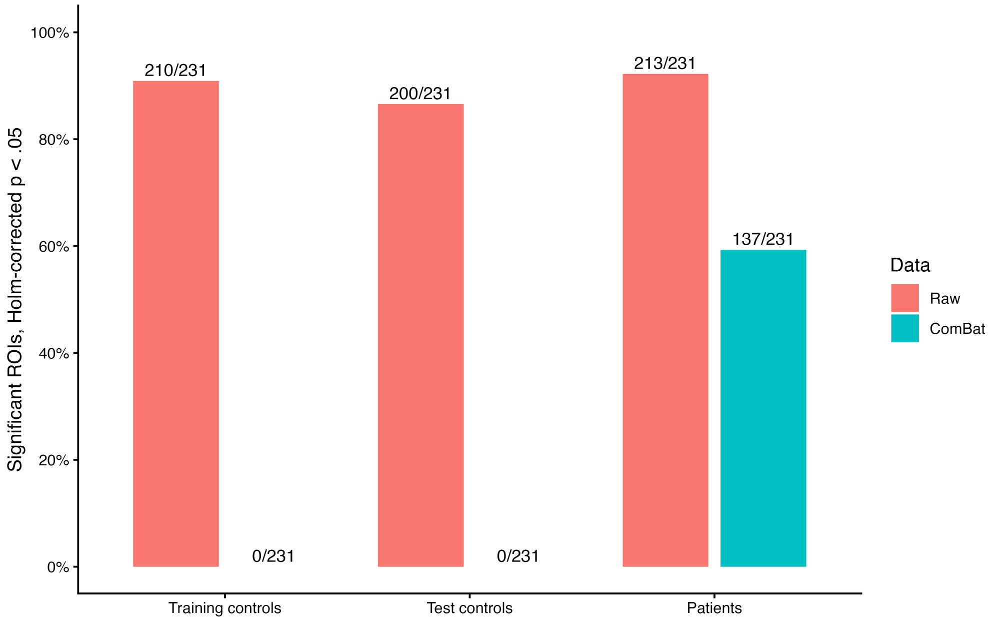


*Note*. Linear ComBat parameters were trained on a HC training cohort and subsequently applied to the unseen $HC_{test}$ cohort and the unseen SSD sample. Raw data is shown in red, ComBat harmonized data is shown in blue. Numbers refer to regions with statistically significant effects of site after Bonferroni-Holm correction in the ANOVAs.

As an additional check of site effects before and after ComBat harmonization, we visually inspected the association between age and regional gray matter volume separated by study. For each gray matter ROI, scatterplots were generated with age on the x-axis and ROI volume on the y-axis. Study-specific linear regression lines with 95% confidence intervals were overlaid to visualize whether the age-volume association differed systematically between sites. This visual inspection was performed separately for the raw and ComBat-harmonized data in the HC training dataset, the independent HC test dataset, and the patient dataset. The visual findings were consistent with the formal ANOVA results, as illustrated in Suppl. Figure S3.

**Suppl. Figure S3**. Age-dependent GMV in an example region before and after harmonization separated by datasets and study

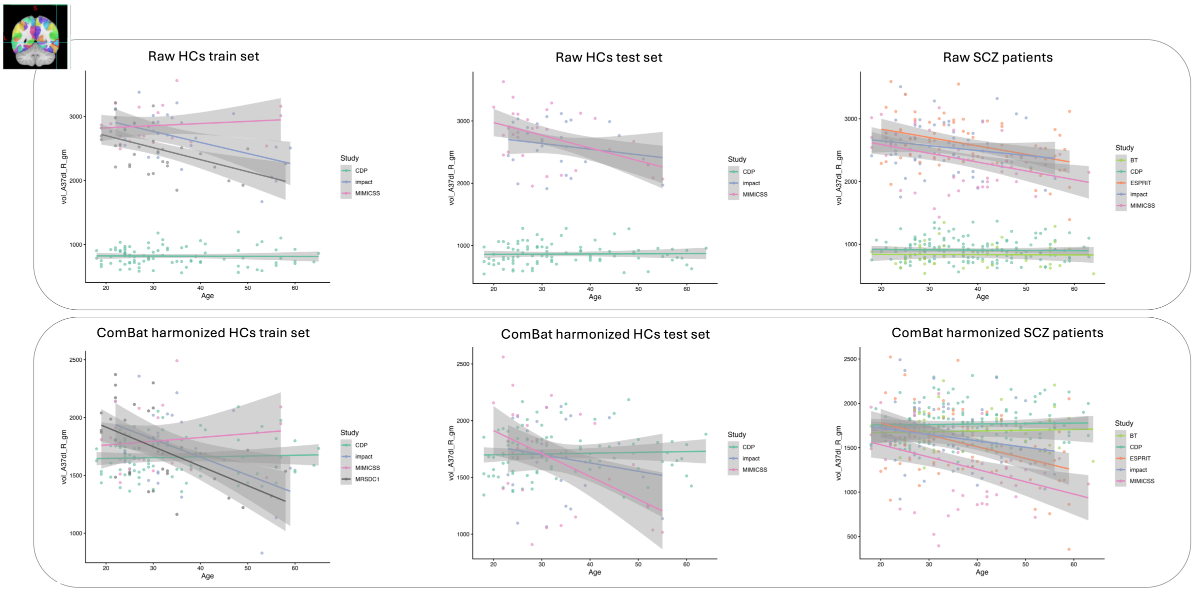


*Note*. Linear ComBat parameters were trained on a HC training cohort and subsequently applied to the unseen HC test cohort and the unseen SSD sample. Slopes represent age-dependent GMV in a dorsolateral area of the middle temporal gyrus. Colors represent different studies. Note that CDP and BT subjects were scanned at the same scanner with the same protocol and shared HC cohorts and ESPRIT and MIMICSS were scanned at the same scanner with the same protocols and shared HC cohorts. The MRSDC1 included only controls, scanned at the same scanner as ESPRIT and MIMICSS.

### Measures

#### *Positive and Negative Syndrome Scale (PANSS[32])*

The PANSS is the gold standard for quantifying psychopathology in patients with schizophrenia. It is a clinician interview comprising two 7-item subscales for negative and positive symptoms, and a general psychopathology subscale with 16 items. All items are rated on a scale from 1 (not existent) to 7 (extreme). A total score is calculated by adding up all items, with a minimum and maximum value of 30 and 210, respectively (7–49 for the positive and negative subscales, 16–112 for the general psychopathology scale).

#### *Trail Making Test Version-B (TMT-B[33])*

In the TMT-B, subjects are instructed to connect numbers and letters alternately in the following order: 1-A-2-B-....-L-13. The time needed in seconds is measured.

### Machine learning classification: SSD vs. HC

We tested the predictive performance of different algorithms, the Least Absolute Shrinkage and Selection Operator (LASSO[34]), Support Vector Machine (SVM[35]), Random Forest (RF[36]) in a nested cross-validation (CV) resampling design. To attain an optimal trade-off between model bias and variance, LASSO classification was included, with $\lambda$ selected by an inner 10-fold CV using the *cv.glmnet* function from the *glmnet* package[37]. For the SVM, the default tuning spaces[38] were used, tuning for the *kernel*, *cost*, *gamma*, and *degree* parameters. A random search (RS[39]) was chosen as a search algorithm, since it has been shown to yield superior performance in higher-dimensional hyperparameter optimization settings compared to grid search[38]. The RS was set to terminate after 100 evaluated parameter configurations, tuning for the area under the curve (AUC). The search space for the *kernel* parameter encompasses a *linear*, *radial*, *sigmoid*, or *polynomial* function. For the latter, the *degree* of the polynomial was tuned in a range from 2 to 5. The *gamma* and *cost* parameters were tuned on a logarithmic scale, with search spaces ranging from 0.0001 to 10,000 (lower and upper limits refer to a linear scale). For the RF, automatic hyperparameter tuning was used to find values for the *mtry* and *sample size* parameter within search spaces from 0 (use no features) to 1 (use all features) and 0.1 (use 10 % of the observations) to 1 (use 100 % of the observations), respectively. Further, as included in the *mlr3* default tuning spaces, the *replace* parameter was tuned, determining whether observations should be drawn from the population with replacement, i.e., using bootstrap samples, or not. To reduce computational efforts due to tuning an additional hyperparameter, the number of trees was set to 1,000, which is slightly higher than the optimal default value[40]. Like for SVMs, a RS tuning for the AUC terminating after 100 evaluations was used as a search algorithm. The out-of-sample performance of the algorithms was evaluated with a 5 × repeated, nested 5-fold CV with 2 folds on the inner loop. Nested resampling is needed to separate model evaluation from model development, including steps of pre-processing and hyperparameter tuning. The final performance estimate for an algorithm is the mean performance averaged across all outer 5 x 5 = 25 CV folds. All model building procedures, including pre-processing and automatic hyperparameter tuning, are implemented in a pipeline (see *mlr3pipelines*[41]) operating in an inner nested resampling loop for performance evaluation. The benchmarking design ensured that all algorithms were trained and evaluated on the same folds, ensuring comparability of the results.

**Suppl. Table S4.** Region-to-atlas-mapping

| Network | Regions |
|---|---|
| AntMTL | vol_A20il_R, vol_A20iv_L, vol_A20iv_R, vol_A20r_L, vol_A20r_R, vol_A20rv_L, vol_A20rv_R, vol_A21r_R, vol_A28_34_L, vol_A35_36c_L, vol_A35_36r_L, vol_A35_36r_R, vol_A38l_L, vol_A38m_L, vol_A38m_R, vol_TI_L, vol_TI_R, vol_lAmyg_L, vol_lAmyg_R, vol_mAmyg_L, vol_mAmyg_R |

| | |
|---|---|
| Auditory | vol_A1_2_3tonIa_L, vol_A1_2_3tonIa_R, vol_A22c_R, vol_A40rv_L, vol_A40rv_R, vol_A41_42_L, vol_A41_42_R, vol_G_L, vol_G_R, vol_TE1.0_TE1.2_L, vol_TE1.0_TE1.2_R, vol_dIg_L, vol_dIg_R |
| CingOperc | vol_A24cd_L, vol_A24cd_R, vol_A40c_R, vol_A44op_L, vol_A44op_R, vol_A44v_L, vol_A44v_R, vol_A4tl_L, vol_A4tl_R, vol_A5m_L, vol_A6m_L, vol_A6m_R, vol_A9_46d_R, vol_dIa_R, vol_dId_L, vol_dId_R, vol_vId_vIg_L, vol_vId_vIg_R |
| Context | vol_A23v_L, vol_A23v_R, vol_A39c_L, vol_A39c_R, vol_TH_L, vol_TH_R, vol_TL_L, vol_TL_R |
| Default | vol_A10l_L, vol_A10m_L, vol_A10m_R, vol_A11m_L, vol_A11m_R, vol_A13_L, vol_A13_R, vol_A14m_L, vol_A14m_R, vol_A20il_L, vol_A21c_L, vol_A21c_R, vol_A21r_L, vol_A31_L, vol_A31_R, vol_A32sg_L, vol_A35_36c_R, vol_A39rv_L, vol_A8dl_L, vol_A8dl_R, vol_A9l_L, vol_A9l_R, vol_aSTS_L |
| DorsAttn | vol_A37elv_R, vol_A37lv_L, vol_A37lv_R, vol_A37vl_R, vol_A6cvl_L, vol_A6cvl_R, vol_A6dl_R, vol_A6vl_L, vol_A7c_L, vol_A7c_R, vol_A7ip_L, vol_A7ip_R, vol_V5_MT+_L, vol_V5_MT+_R, vol_lsOccG_L, vol_lsOccG_R |
| FaceSM | vol_A1_2_3ulhf_L, vol_A1_2_3ulhf_R, vol_A4hf_L, vol_A4hf_R |
| FootSM | vol_A1_2_3ll_L, vol_A1_2_3ll_R, vol_A1_2_3tru_L, vol_A1_2_3tru_R, vol_A4ll_L, vol_A4ll_R, vol_A4t_L, vol_A4t_R, vol_A4ul_L |
| FrontPar | vol_A10l_R, vol_A11l_L, vol_A11l_R, vol_A20cl_L, vol_A20cl_R, vol_A20cv_L, vol_A20cv_R, vol_A37elv_L, vol_A37vl_L, vol_A39rd_L, vol_A39rd_R, vol_A40c_L, vol_A44d_L, vol_A44d_R, vol_A6vl_R, vol_A8vl_L, vol_A8vl_R, vol_A9_46v_L, vol_A9_46v_R, vol_IFJ_L, vol_IFJ_R, vol_IFS_L, vol_IFS_R |
| HandSM | vol_A2_L, vol_A2_R, vol_A4ul_R, vol_A6cdl_L, vol_A6cdl_R |
| Language | vol_A22c_L, vol_A22r_L, vol_A22r_R, vol_A37dl_L, vol_A37dl_R, vol_A38l_R, vol_A39rv_R, vol_A45c_L, vol_A45c_R, vol_A45r_L, vol_A45r_R, vol_A8m_L, vol_A8m_R, vol_aSTS_R, vol_cpSTS_L, vol_cpSTS_R, vol_rpSTS_L, vol_rpSTS_R |
| LatVis | vol_A37mv_L, vol_A37mv_R, vol_OPC_L, vol_OPC_R, vol_cLinG_L, vol_cLinG_R, vol_iOccG_L, vol_iOccG_R, vol_mOccG_L, vol_mOccG_R, vol_msOccG_L, vol_msOccG_R, vol_rLinG_L, vol_rLinG_R |
| MedVis | vol_cCunG_L, vol_cCunG_R, vol_rCunG_L, vol_rCunG_R, vol_vmPOS_L, vol_vmPOS_R |

| | |
|---|---|
| ParMemory | vol_A23c_L, vol_A23c_R, vol_A23d_L, vol_A23d_R, vol_A24rv_L, vol_A7m_L, vol_A7m_R, vol_dmPOS_L, vol_dmPOS_R |
| PostMTL | vol_A28_34_R, vol_cHipp_L, vol_cHipp_R, vol_rHipp_L, vol_rHipp_R |
| Premotor | vol_A40rd_L, vol_A40rd_R, vol_A5l_L, vol_A5l_R, vol_A5m_R, vol_A6dl_L, vol_A7pc_L, vol_A7pc_R, vol_A7r_L, vol_A7r_R |
| Salience | vol_A12_47l_L, vol_A12_47l_R, vol_A12_47o_L, vol_A12_47o_R, vol_A24rv_R, vol_A32p_L, vol_A32p_R, vol_A32sg_R, vol_A46_L, vol_A46_R, vol_A9_46d_L, vol_A9m_L, vol_A9m_R, vol_dIa_L, vol_vIa_L, vol_vIa_R |

*Note*. Brainnetome atlas regions are assigned to networks based on the highest dice coefficient (Network Correspondence Toolbox[42]). AntMTL: Anterior Medial Temporal Lobe. CingOperc: Cingulo-Opercular. DorsAttn: Dorsal Attention. SM: Somatomotor.

### Supplementary Material 2: Results

**Suppl. Figure S4.** PANSS scores for patients, separated by study

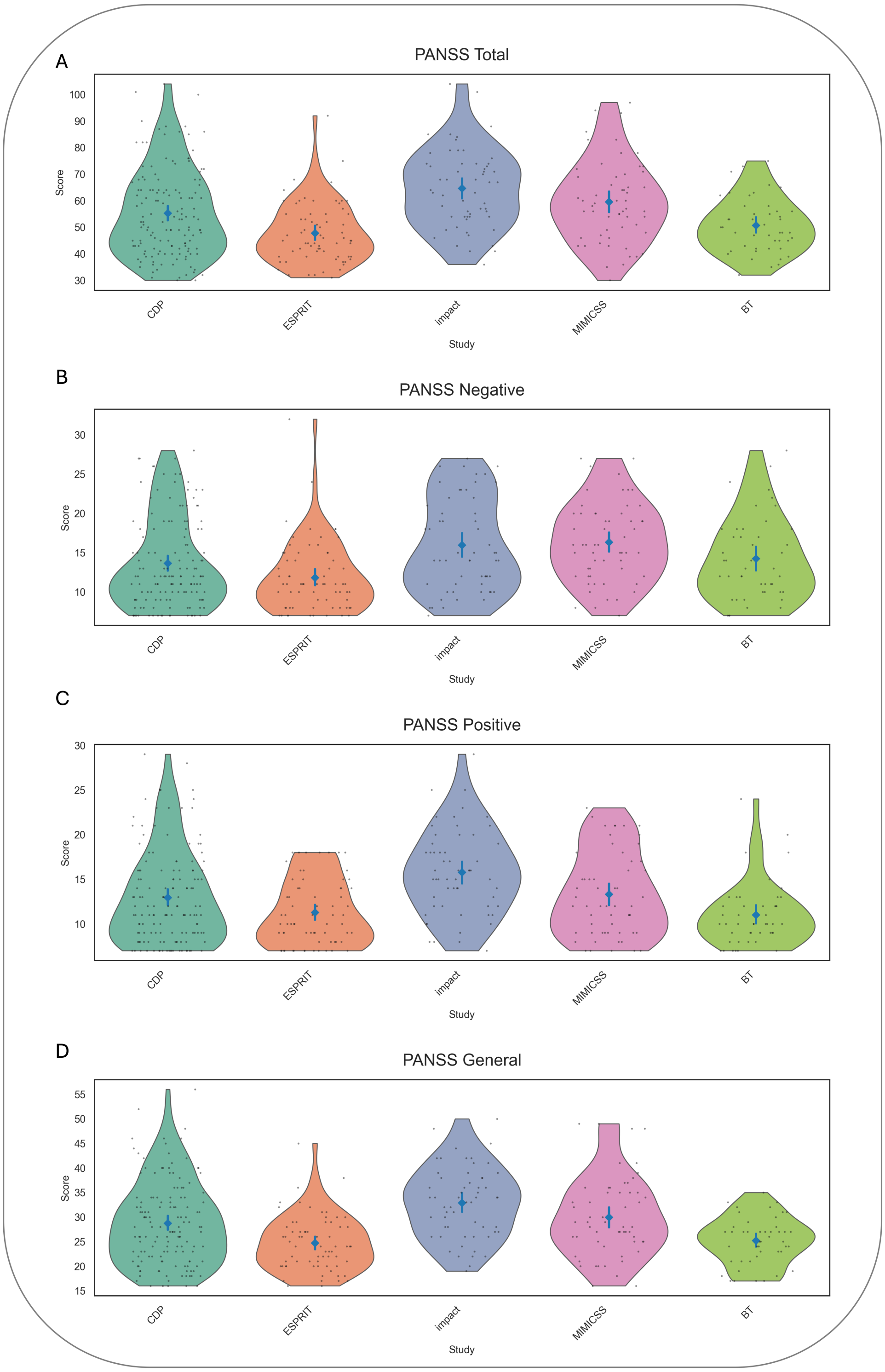


*Note*. PANSS total and subscale scores separated for the clinical studies. PANSS: Positive and Negative Syndrome Scale. CDP: Clinical Deep Phenotyping. ESPRIT: Enhancing Schizophrenia Prevention and Recovery through Innovative Treatments. MIMICSS: Multimodal Imaging in Chronic Schizophrenia Study. BT: BrainTrain.

**Suppl. Figure S5.** Mean Z-scores separated by study

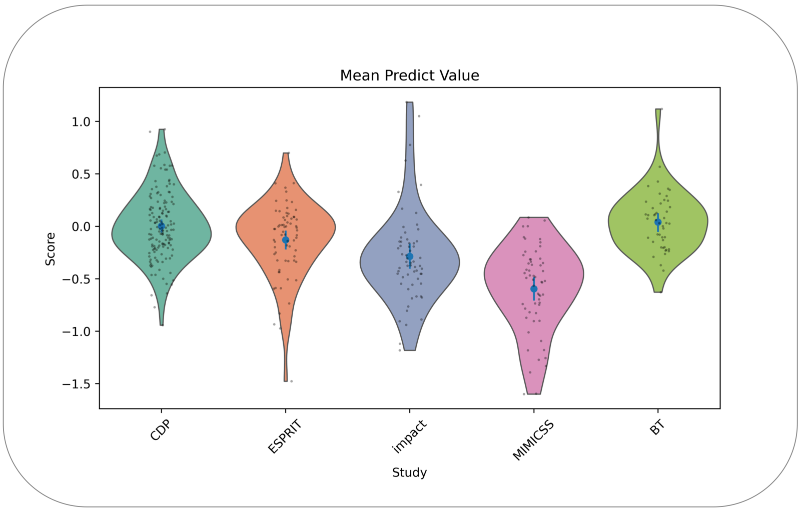


*Note*. Mean Z-scores separated for the clinical studies. CDP: Clinical Deep Phenotyping. ESPRIT: Enhancing Schizophrenia Prevention and Recovery through Innovative Treatments. MIMICSS: Multimodal Imaging in Chronic Schizophrenia Study. BT: BrainTrain

**Suppl. Figure S6**. Model fit metrics

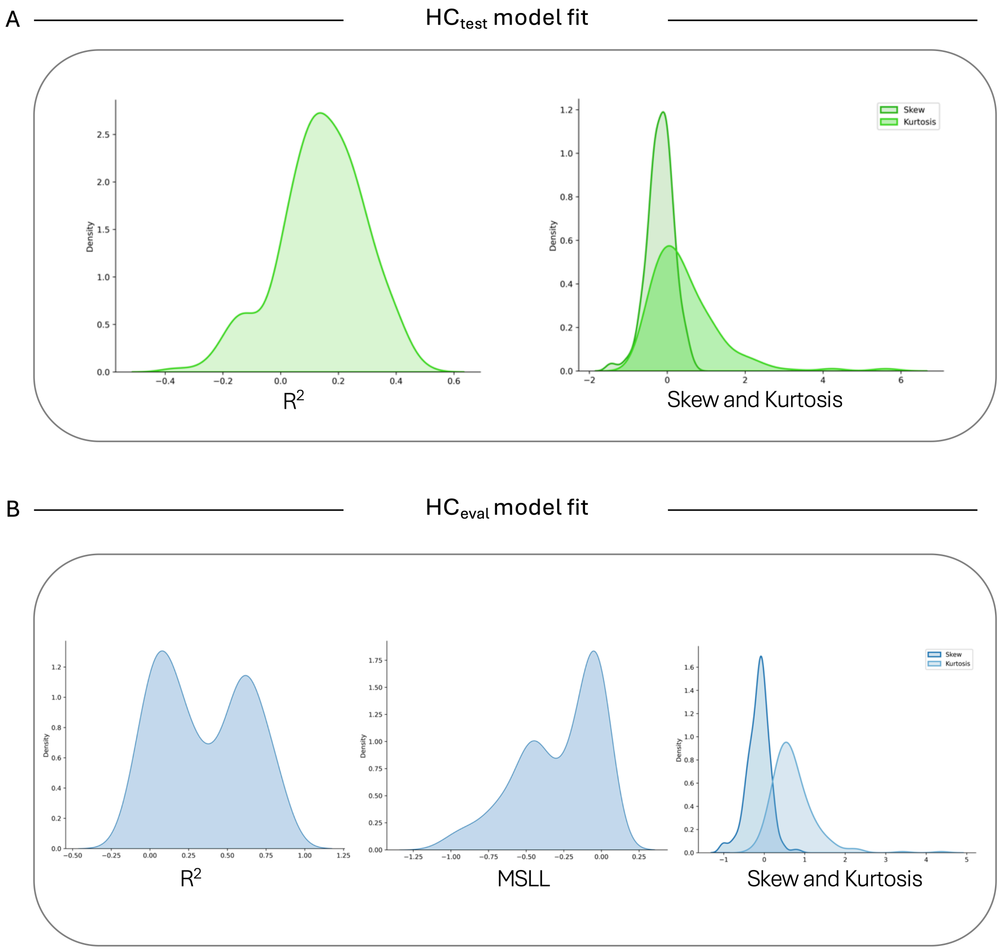

*Note*. Panel A: R squared (left) and skew and kurtosis (right) for the $HC_{test}$ sample. Panel B: R squared (left), Mean Standardized Log Loss (MSLL), skew and kurtosis (right) for the $HC_{eval}$ sample.

**Suppl. Table S5.** Out-of-sample Performances for Classification Models of HCs vs. SSD

| algorithm | *AUC* | | *BAC* | | *SENS* | *SPEC* | *BS* |
|---|---|---|---|---|---|---|---|
| | *M* | *SD* | *M* | *SD* | | | |
| Featureless learner | 0.50 | 0.00 | 0.50 | 0.00 | 1.00 | 0.00 | 0.20 |
| LASSO regression | 0.75 (0.78) | 0.04 (0.03) | 0.54 (0.72) | 0.03 (0.04) | 0.97 (0.75) | 0.10 (0.69) | 0.18 (0.19) |
| Random Forest | 0.77 (0.81) | 0.05 (0.03) | 0.56 (0.73) | 0.04 (0.03) | 0.97 (0.77) | 0.16 (0.69) | 0.17 (0.19) |
| Support Vector machine | 0.79 (0.83) | 0.05 (0.03) | 0.63 (0.75) | 0.07 (0.03) | 0.92 (0.80) | 0.33 (0.71) | 0.17 (0.17) |

*Note.* N = 149 for HCs and n = 379 for SSD patients. Out-of-sample performance is estimated with a 5 × repeated 5-fold nested cross-validation with 2 folds on the inner loop. M represents the mean, and SD represents the standard deviation of performance estimates across the outer resampling folds. Values in parentheses refer to performance estimates when leveraging the calibration dataset for the HC cohort, leading to an increased sample of n = 324 for HCs. *BAC* = Balanced accuracy; *BS* = Brier Score; *SENS* = Sensitivity; *SPEC* = Specificity; *AUC* = Area under the curve; *LASSO* = Least Absolute Shrinkage and Selection Operator. HC: Healthy control. SSD: Schizophrenia Spectrum Disorder.

**Suppl. Table S6.** Raw data out-of-sample performances for Classification Models of HCs vs. SSD

| algorithm | *AUC* | | *BAC* | | *SENS* | *SPEC* | *BS* |
|---|---|---|---|---|---|---|---|
| | *M* | *SD* | *M* | *SD* | | | |
| Featureless learner | 0.50 | 0.00 | 0.50 | 0.00 | 1.00 | 0.00 | 0.20 |
| LASSO regression | 0.76 | 0.05 | 0.54 | 0.04 | 0.97 | 0.11 | 0.18 |
| Random Forest | 0.76 | 0.05 | 0.58 | 0.03 | 0.96 | 0.19 | 0.17 |
| Support Vector machine | 0.77 | 0.05 | 0.61 | 0.06 | 0.92 | 0.30 | 0.17 |

*Note.* N = 149 for HCs and n = 379 for SSD patients. Out-of-sample performance is estimated with a 5 × repeated 5-fold nested cross-validation with 2 folds on the inner loop. M represents the mean, and SD represents the standard deviation of performance estimates across the outer resampling folds. *BAC* = Balanced accuracy; *BS* = Brier Score; *SENS* = Sensitivity;

*SPEC* = Specificity; *AUC* = Area under the curve; *LASSO* = Least Absolute Shrinkage and Selection Operator. HC: Healthy control. SSD: Schizophrenia Spectrum Disorder.

**Suppl. Table S7**. Network-specific ADS

| | mean $ADS_{Network}$ | |
|---|---|---|
| functional network | SSD | HC |
| Salience | -0.3379 | -0.0386 |
| Default | -0.2738 | -0.0457 |
| Language | -0.2716 | -0.0227 |
| CingOperc | -0.2651 | -0.0242 |
| AntMTL | -0.2582 | 0.0201 |
| FrontPar | -0.2548 | -0.0344 |
| ParMemory | -0.2325 | -0.0033 |
| Auditory | -0.2087 | -0.0045 |
| DorsAttn | -0.1882 | 0.0183 |
| PostMTL | -0.1816 | -0.0196 |
| HandSM | -0.1795 | -0.0010 |
| Premotor | -0.1730 | -0.0472 |
| Context | -0.1665 | 0.0090 |
| FootSM | -0.0854 | 0.0156 |
| FaceSM | -0.0612 | 0.0290 |
| LatVis | -0.0343 | 0.0442 |
| MedVis | -0.0256 | -0.0729 |

*Note*. Average deviation scores (ADS) for functional brain networks in the patient and control group. $N_{SSD}$ = 264; $N_{HC}$ = 149. Network-level analyses were restricted to clinical studies that included both SSD patients and HCs (CDP, IMPACT, MIMICSS). SSD: schizophrenia spectrum disorder. HC: healthy control.

**Suppl. Table S8**. Regression parameters for network-specific analyses.

| | group difference | | | group difference controlling for global network deviation | | |
|---|---|---|---|---|---|---|
| functional network | Estimate | *SE* | *p* | Estimate | *SE* | *p* |
| Salience | -0.299 | 0.058 | <.001 | -0.096 | 0.028 | <.001 |
| | (-0.284) | (0.054) | (<.001) | (-0.097) | (0.028) | (<.001) |
| Default | -0.228 | 0.058 | <.001 | -0.025 | 0.028 | .359 |
| | (-0.210) | (0.052) | (<.001) | (-0.028) | (0.028) | (.321) |
| Language | -0.249 | 0.052 | <.001 | -0.065 | 0.024 | .007 |
| | (-0.230) | (0.046) | (<.001) | (-0.067) | (0.024) | (.006) |
| CingOperc | -0.241 | 0.049 | <.001 | -0.068 | 0.022 | .002 |
| | (-0.229) | (0.045) | (<.001) | (-0.069) | (0.022) | (.002) |
| AntMTL | -0.278 | 0.047 | <.001 | -0.154 | 0.036 | <.001 |
| | (-0.271) | (0.045) | (<.001) | (-0.153) | (0.036) | (<.001) |
| FrontPar | -0.220 | 0.052 | <.001 | -0.037 | 0.024 | .118 |
| | (-0.206) | (0.047) | (<.001) | (-0.039) | (0.024) | (.099) |
| ParMemory | -0.229 | 0.055 | <.001 | -0.056 | 0.034 | .105 |
| | (-0.205) | (0.051) | (<.001) | (-0.045) | (0.033) | (.172) |
| Auditory | -0.204 | 0.054 | <.001 | -0.024 | 0.030 | .412 |
| | (-0.188) | (0.048) | (<.001) | (-0.032) | (0.029) | (.277) |
| DorsAttn | -0.206 | 0.050 | <.001 | -0.032 | 0.024 | .183 |
| | (-0.190) | (0.046) | (<.001) | (-0.032) | (0.024) | (.193) |
| PostMTL | -0.162 | 0.059 | .007 | -0.012 | 0.047 | .799 |
| | (-0.144) | (0.057) | (.013) | (0.000) | (0.046) | (.999) |
| HandSM | -0.179 | 0.064 | .006 | 0.036 | 0.036 | .318 |
| | (-0.163) | (0.060) | (.007) | (0.037) | (0.036) | (.309) |
| Premotor | -0.126 | 0.058 | .031 | 0.062 | 0.035 | .073 |
| | (-0.117) | (0.055) | (.035) | (0.063) | (0.034) | (.064) |
| Context | -0.176 | 0.054 | .001 | -0.010 | 0.036 | .772 |
| | (-0.151) | (0.049) | (.002) | (-0.010) | (0.035) | (.782) |
| FootSM | -0.101 | 0.064 | .117 | 0.110 | 0.037 | .003 |
| | (-0.088) | (0.060) | (.146) | (0.108) | (0.037) | (.004) |
| FaceSM | -0.090 | 0.072 | .210 | 0.138 | 0.044 | .002 |
| | (-0.073) | (0.065) | (.262) | (0.126) | (0.044) | (.004) |

| | | | | | | |
|---|---|---|---|---|---|---|
| LatVis | -0.079 (-0.058) | 0.056 (0.052) | .163 (.263) | 0.075 (0.074) | 0.042 (0.041) | .072 (.075) |
| MedVis | 0.047 (0.055) | 0.044 (0.043) | .288 (.203) | 0.159 (0.162) | 0.035 (0.035) | <.001 (<.001) |

*Note.* $N_{SSD}$ = 264; $N_{HC}$ = 149. Network-level analyses were restricted to clinical studies that included both SSD patients and HCs (CDP, IMPACT, MIMICSS). HCs are coded as the reference category. Estimates are unstandardized regression coefficients for the group factor. Values in parentheses indicate group regression parameters and p-values when additionally controlling for study (sensitivity analysis). HC: healthy control. SSD: schizophrenia spectrum disorder.

**Sensitivity analyses**

First, to test the robustness of the results against cutoff criteria, we repeated the outlier quality control by only excluding the manually checked data artefacts, refraining from cutoff criteria above which values were set to NA in the original analyses. The confirmatory and exploratory group comparisons and correlation analyses were repeated analogously. Overall, we found highly comparable results to the original analyses.

For H1, we compared average deviation scores (ADS) between SSD vs. HCs, assuming a greater negative deviation in patients. We found a significantly lower ADS in SSD ($t(526) = -3.90$, one-sided $p < .001$) with an effect size of $d = -0.38$. Comparing the percentages of extreme negative Z-scores as a secondary outcome measure, we found a significantly higher extreme negative value percentage in the SSD compared to the HC group (U = 37702, one-sided $p < .001$; $r = 0.34$).

For H2, we replicated the hypothesized negative Spearman correlation between ADS and PANSS total in the SSD group ($\rho(368) = -0.18$; 95% CI [−0.28, −0.08]; one-sided $p < .001$) and exploratorily for the PANSS negative ($\rho(369) = -0.19$; 95% CI [−0.29, −0.09]; one-sided p (r < 0) < 0.001) and PANSS general ($\rho(369) = -0.16$; 95% CI [−0.26, −0.06]; one-sided $p < .001$).

For H3, we found the hypothesized negative Spearman correlation between the ADS and the TMT-B in the SSD group ($\rho(264) = -0.23$; 95% CI [−0.34, −0.11]; one-sided $p < .001$).

Second, we repeated confirmatory and exploratory analyses using only data from the CDP study to refrain from data harmonization between studies. As the imputation procedure was applied to harmonized data, no imputation was performed in this analysis. For H1, comparing ADS between SSD vs. HCs, we found a significantly lower ADS in SSD ($t(141) = -2.64$, one-sided $p = .005$). For H2, we found a negative Spearman correlation between the ADS and PANSS total in the SSD sample ($\rho(86) = -0.16$; 95% CI [−0.36, 0.05]; one-sided $p = .070$). The ADS was negatively associated with the PANSS negative subscale ($\rho(86) = -0.15$; 95% CI [−0.35, 0.06]; one-sided $p = .079$) and the PANSS general ($\rho(86) = -0.17$; 95% CI [−0.37, 0.04];

one-sided $p = .056$). For H3, we found a negative Spearman correlation between the ADS and the TMT-B ($\rho(23) = -0.27$; 95% CI [−0.60; 0.14]; one-sided $p = .094$). Overall, despite H2 and H3 not reaching statistical significance (which is expected due to the reduced sample size), the direction and size of the effect remained comparable to the collapsed study sample.

**Supplementary analysis**

***Raw features***

We correlated the mean raw GMV with the PANSS scores controlling for eTIV, age and sex in the sample. All Spearman correlation coefficients were non-significant (PANSS total: $\rho(365) = -0.02$, 95% CI [−0.12, 0.09]; one-sided $p = .371$; PANSS negative: $\rho(366) = -0.07$, 95% CI [−0.17, 0.03], one-sided $p = .085$; PANSS general: $\rho(366) = 0.00$, 95% CI [−0.11, 0.10], one-sided $p = .463$). For cognition, the negative correlation remained significant at an uncorrected significance level (TMT-B; $\rho(261) = -0.23$; 95% CI [−0.34, −0.11]; one-sided $p < .001$).

***Effects of medication, illness duration and body mass index (BMI) on deviation scores***

There was a significant negative Spearman correlation between the ADS and chlorpromazine equivalents (CPZ) in the SSD group ($\rho(355) = -0.15$; 95% CI [−0.25, −0.04]; $p = .006$). Illness duration was significantly positively correlated with the ADS ($\rho(371) = 0.13$; 95% CI [0.02, 0.22]; $p = .015$). There was no significant Pearson correlation between the ADS and BMI in the SSD group ($r(268) = 0.01$; 95% CI [−0.11, 0.13]; $p = .831$).

**Consortia**

***BrainTrain Working Group***. Marina Amann, Jakob Bauereiß, Lena Deller, Jonathan Diegelmann, Peter Falkai, Josephine Glad, Mona Hussain, Jasmin Jannan, Miriam John, Oezlem Koç, Susanne Kuprat, Klara Lada, Isabel Maurus, Joanna Moussiopoulou, Susanne Münz, Lukas Roell, Linda Sagstetter, Jana Sautner, Andrea Schmitt, Julia Segerer, Johanna Spaeth, Nina Theis, Julia Graefin von Wartensleben, Annemarie Weibel, Lara Widmann, Vladislav Yakimov, Devin Yildirim, Deniz Yilmaz, Miriam Zuliani.

***CDP Working Group.*** Stephanie Behrens, Emanuel Boudriot, Man-Hsin Chang, Valéria de Almeida, Sylvia de Jonge, Fanny Dengl, Peter Falkai, Laura E. Fischer, Nadja Gabellini, Vanessa Gabriel, Sabrina Galinski, Thomas Geyer, Katharina Hanken, Alkomiet Hasan, Genc Hasanaj, Alexandra Hisch, Georgios Ioannou, Iris Jäger, Marcel Kallweit, Temmuz Karali, Susanne Karch, Berkhan Karslı, Daniel Keeser, Christoph Kern, Nicole L. Klimas, Maxim Korman, Nikolaos Koutsouleris, Lenka Krcmar, Verena Meisinger, Julian Melcher, Matin Mortazavi, Joanna Moussiopoulou, Karin Neumeier, Frank Padberg, Boris Papazov, Irina Papazova, Sergi Papiol, Pauline Pingen, Oliver Pogarell, Siegfried G. Priglinger, Florian J. Raabe, Lukas Roell, Moritz J. Rossner, Philipp Sämann, Andrea Schmitt, Susanne

Schmölz, Eva C. Schulte, Enrico Schulz, Benedikt Schworm, Zhuanghua Shi, Elias Wagner, Sven Wichert, Vladislav Yakimov, Peter Zill, Michael J. Ziller.

### References

1. Krčmář, L., Jäger, I., Boudriot, E., Hanken, K., Gabriel, V., Melcher, J., *et al.* The multimodal Munich Clinical Deep Phenotyping study to bridge the translational gap in severe mental illness treatment research. *Front. Psychiatry* **14**, 1179811 (2023).
2. Maurus, I., Roell, L., Lembeck, M., Papazova, I., Greska, D., Muenz, S., *et al.* Exercise as an add-on treatment in individuals with schizophrenia: Results from a large multicenter randomized controlled trial. *Psychiatry Res.* **328**, 115480 (2023).
3. Moussiopoulou, J., Yakimov, V., Roell, L., Rauchmann, B.-S., Toth, H., Melcher, J., *et al.* Higher blood–brain barrier leakage in schizophrenia-spectrum disorders: A comparative dynamic contrast-enhanced magnetic resonance imaging study with healthy controls. *Brain. Behav. Immun.* **128**, 256–265 (2025).
4. Sheehan, D. V., Lecrubier, Y., Sheehan, K. H., Amorim, P., Janavs, J., Weiller, E., *et al.* The Mini-International Neuropsychiatric Interview (M.I.N.I.): the development and validation of a structured diagnostic psychiatric interview for DSM-IV and ICD-10. *J. Clin. Psychiatry* **59 Suppl 20**, 22-33;quiz 34-57 (1998).
5. Budde, M., Anderson-Schmidt, H., Gade, K., Reich-Erkelenz, D., Adorjan, K., Kalman, J. L., *et al.* A longitudinal approach to biological psychiatric research: The PsyCourse study. *Am. J. Med. Genet. Part B Neuropsychiatr. Genet. Off. Publ. Int. Soc. Psychiatr. Genet.* **180**, 89–102 (2019).
6. Maurus, I., Hasan, A., Schmitt, A., Roeh, A., Keeser, D., Malchow, B., *et al.* Aerobic endurance training to improve cognition and enhance recovery in schizophrenia: design and methodology of a multicenter randomized controlled trial. *Eur. Arch. Psychiatry Clin. Neurosci.* **271**, 315–324 (2021).
7. Keeser, D., Meindl, T., Bor, J., Palm, U., Pogarell, O., Mulert, C., *et al.* Prefrontal transcranial direct current stimulation changes connectivity of resting-state networks during fMRI. *J. Neurosci. Off. J. Soc. Neurosci.* **31**, 15284–15293 (2011).

8. Shafto, M. A., Tyler, L. K., Dixon, M., Taylor, J. R., Rowe, J. B., Cusack, R., *et al.* The Cambridge Centre for Ageing and Neuroscience (Cam-CAN) study protocol: a cross-sectional, lifespan, multidisciplinary examination of healthy cognitive ageing. *BMC Neurol.* **14**, 204 (2014).

9. Taylor, J. R., Williams, N., Cusack, R., Auer, T., Shafto, M. A., Dixon, M., *et al.* The Cambridge Centre for Ageing and Neuroscience (Cam-CAN) data repository: Structural and functional MRI, MEG, and cognitive data from a cross-sectional adult lifespan sample. *NeuroImage* **144**, 262–269 (2017).

10. Miller, K. L., Alfaro-Almagro, F., Bangerter, N. K., Thomas, D. L., Yacoub, E., Xu, J., *et al.* Multimodal population brain imaging in the UK Biobank prospective epidemiological study. *Nat. Neurosci.* **19**, 1523–1536 (2016).

11. Sudlow, C., Gallacher, J., Allen, N., Beral, V., Burton, P., Danesh, J., *et al.* UK Biobank: An Open Access Resource for Identifying the Causes of a Wide Range of Complex Diseases of Middle and Old Age. *PLOS Med.* **12**, e1001779 (2015).

12. Van Essen, D. C., Smith, S. M., Barch, D. M., Behrens, T. E. J., Yacoub, E. & Ugurbil, K. The WU-Minn Human Connectome Project: An Overview. *NeuroImage* **80**, 62–79 (2013).

13. Harms, M. P., Somerville, L. H., Ances, B. M., Andersson, J., Barch, D. M., Bastiani, M., *et al.* Extending the Human Connectome Project across ages: Imaging protocols for the Lifespan Development and Aging projects. *NeuroImage* **183**, 972–984 (2018).

14. Bookheimer, S. Y., Salat, D. H., Terpstra, M., Ances, B. M., Barch, D. M., Buckner, R. L., *et al.* The Lifespan Human Connectome Project in Aging: An overview. *NeuroImage* **185**, 335–348 (2019).

15. Horta, M., Polk, R. & Ebner, N. Single Dose Intranasal Oxytocin Administration: Data from Healthy Younger and Older Adults. Openneuro https://doi.org/10.18112/OPENNEURO.DS004725.V1.0.1 (2023).

16. Tisdall, L. & Mata, R. Age differences in the neural basis of decision-making under uncertainty. *Cogn. Affect. Behav. Neurosci.* **23**, 788–808 (2023).

17. Rogers, Jones, McConkey & Peelle. Listening task. OpenNeuro https://doi.org/10.18112/openneuro.ds004285.v1.0.0 (2022).
18. Nugent, Thomas, Mahoney, Gibbons, Smith, Charles, *et al.* The NIMH Healthy Research Volunteer Dataset. OpenNeuro https://doi.org/10.18112/openneuro.ds004215.v1.0.3 (2024).
19. Fialkowski, K., Messias, I. & Bush, K. A. Cognitive Control Theoretic Mechanisms of Real-time fMRI-Guided Neuromodulation (CTM). Openneuro https://doi.org/10.18112/OPENNEURO.DS003831.V1.0.0 (2021).
20. Bush, K. A., James, G. A., Privratsky, A. A., Fialkowski, K. P. & Kilts, C. D. Action-value processing underlies the role of the dorsal anterior cingulate cortex in performance monitoring during self-regulation of affect. *PLOS ONE* **17**, e0273376 (2022).
21. Van Der Laan, L. N., Scholz, C., Poldrack, R. A., De Ridder, D. T. D. & Smidts, A. Can we have a second helping? A replication study on the neurobiological mechanisms underlying self-control. Openneuro https://doi.org/10.18112/OPENNEURO.DS002643.V1.1.0 (2022).
22. Nastase, S. A., Liu, Y.-F., Hillman, H., Asieh Zadbood, Hasenfratz, L., Neggin Keshavarzian, *et al.* Narratives. Openneuro https://doi.org/10.18112/OPENNEURO.DS002345.V1.1.4 (2020).
23. Champely, S., Ekstrom, C., Dalgaard, P., Gill, J., Weibelzahl, S., Anandkumar, A., *et al.* pwr: Basic Functions for Power Analysis. (2020).
24. Bethlehem, R. A. I., Seidlitz, J., White, S. R., Vogel, J. W., Anderson, K. M., Adamson, C., *et al.* Brain charts for the human lifespan. *Nature* **604**, 525–533 (2022).
25. Bayer, J. M. M., Dinga, R., Kia, S. M., Kottaram, A. R., Wolfers, T., Lv, J., *et al.* Accommodating site variation in neuroimaging data using normative and hierarchical Bayesian models. *NeuroImage* **264**, 119699 (2022).

26. Esteban, O., Birman, D., Schaer, M., Koyejo, O. O., Poldrack, R. A. & Gorgolewski, K. J. MRIQC: Advancing the automatic prediction of image quality in MRI from unseen sites. *PLOS ONE* **12**, e0184661 (2017).
27. Young, T. R., Kumar, V. J. & Saranathan, M. Normative Modeling of Thalamic Nuclear Volumes and Characterization of Lateralized Volume Alterations in Alzheimer's Disease Versus Schizophrenia. *Biol. Psychiatry Cogn. Neurosci. Neuroimaging* **10**, 726–739 (2025).
28. Albu, E., Gao, S., Wynants, L. & Van Calster, B. missForestPredict—Missing data imputation for prediction settings. *PLOS ONE* **20**, e0334125 (2025).
29. Fraza, C. J., Dinga, R., Beckmann, C. F. & Marquand, A. F. Warped Bayesian linear regression for normative modelling of big data. *NeuroImage* **245**, 118715 (2021).
30. Bayer, J. M. M., Thompson, P. M., Ching, C. R. K., Liu, M., Chen, A., Panzenhagen, A. C., *et al.* Site effects how-to and when: An overview of retrospective techniques to accommodate site effects in multi-site neuroimaging analyses. *Front. Neurol.* **13**, 923988 (2022).
31. Radua, J. combat.enigma: Fit and Apply ComBat, LMM, or Prescaling Harmonization for ENIGMA and Other Multisite MRI Data. (2024).
32. Kay, S. R., Fiszbein, A. & Opler, L. A. The positive and negative syndrome scale (PANSS) for schizophrenia. *Schizophr. Bull.* **13**, 261–276 (1987).
33. Reitan, R. M. & Wolfson, D. *The Halstead-Reitan Neuropsychological Test Battery: Theory and Clinical Interpretation*. (Neuropsychology Press, Tucson, Ariz, 1985).
34. Tibshirani, R. Regression Shrinkage and Selection via the Lasso. *J. R. Stat. Soc. Ser. B Methodol.* **58**, 267–288 (1996).
35. Cortes, C. & Vapnik, V. Support-vector networks. *Mach. Learn.* **20**, 273–297 (1995).
36. Breiman, L. Random Forests. *Mach. Learn.* **45**, 5–32 (2001).
37. Friedman, J. H., Hastie, T. & Tibshirani, R. Regularization Paths for Generalized Linear Models via Coordinate Descent. *J. Stat. Softw.* **33**, 1–22 (2010).

38. Bischl, B., Binder, M., Lang, M., Pielok, T., Richter, J., Coors, S., *et al.* Hyperparameter optimization: Foundations, algorithms, best practices, and open challenges. *WIREs Data Min. Knowl. Discov.* **13**, e1484 (2023).
39. Bergstra, J. & Bengio, Y. Random Search for Hyper-Parameter Optimization. *J. Mach. Learn. Res.* **13**, 281–305 (2012).
40. Probst, P., Wright, M. N. & Boulesteix, A.-L. Hyperparameters and tuning strategies for random forest. *WIREs Data Min. Knowl. Discov.* **9**, e1301 (2019).
41. Binder, M., Pfisterer, F., Lang, M., Schneider, L., Kotthoff, L. & Bischl, B. mlr3pipelines - Flexible Machine Learning Pipelines in R. *J. Mach. Learn. Res.* **22**, 1–7 (2021).
42. Kong, R., Spreng, R. N., Xue, A., Betzel, R. F., Cohen, J. R., Damoiseaux, J. S., *et al.* A network correspondence toolbox for quantitative evaluation of novel neuroimaging results. *Nat. Commun.* **16**, 2930 (2025).
